# Integrative multi-omic analyses identify major axes of heterogeneity in chronic obstructive pulmonary disease and uncover their molecular contributors

**DOI:** 10.64898/2026.01.22.26344654

**Authors:** Arda Halu, Matthew Moll, Chengyue Zhang, Leonardo Martini, Per S. Bakke, Russell P. Bowler, Peter J. Castaldi, Michael H. Cho, Dawn L. DeMeo, Kimberly Glass, Craig P. Hersh, Brian D. Hobbs, Edwin K. Silverman

## Abstract

Chronic obstructive pulmonary disease (COPD) is a debilitating and progressive lung disease that affects millions of people worldwide. There is a continuing clinical need to characterize COPD at the molecular level to be able to identify the multi-omic biomarkers of its pathogenesis and to enable more accurate diagnoses and more effective treatment. We used Multi-Omics Factor Analysis (MOFA) to jointly analyze genomic, blood transcriptomic, and plasma proteomic data collected from 1,872 participants in the Genetic Epidemiology of COPD study who had moderate to very severe COPD. Five latent factors identified by MOFA were associated with COPD-related lung function, chest computed tomography (CT) imaging, and blood count phenotypes, as well as all-cause mortality. The top genetic, transcriptomic and proteomic contributors to these latent factors were also individually associated with COPD-related outcomes. Moreover, factor loadings and expression levels of top omic drivers helped distinguish between patient subgroups. Quantitative trait loci analysis of a latent factor that was jointly driven by transcriptomics and proteomics revealed potential common genetic control of gene expression and protein abundance. Polygenic risk scores derived from a genomics-driven latent factor were associated with chest CT imaging and lung function phenotypes, and these associations were replicated in an independent COPD cohort. Together, our results suggest the potential of integrative omic approaches to identify the major axes of heterogeneity in COPD and uncover the multi-omic interplay between the contributors to each axis.

## Introduction

Chronic obstructive pulmonary disease (COPD) is the most prevalent chronic lung disorder worldwide with an enormous morbidity and mortality burden (1,2), and its management is currently limited to treating symptoms, slowing disease progression, and preventing exacerbations (3). Clinically, COPD is a heterogeneous syndrome with high variability between individuals in terms of disease onset and progression, and response to treatment (4,5). The classical phenotypes of “pink puffer” (underweight with emphysema) and “blue bloater” (overweight with chronic bronchitis) are now considered to be reductive, as most patients present with some combination of the many characteristics of these phenotypes (6). Due to this heterogeneity, it has been an ongoing challenge to formulate a precise definition of the disease (7). For a more accurate and precise clinical diagnosis of complex diseases such as COPD, identifying subpopulations of patients with similar clinical and molecular characteristics, a practice known as disease subtyping, is critical.

While numerous clinical subtypes of COPD have been identified via computational methods, most of these approaches attempt to yield discrete clusters, i.e., non-overlapping groups of individuals, which have proven challenging to reproduce across different studies (8). Due to the large number of clinical and molecular contributors to COPD, it is plausible that its clinical subtypes are overlapping at least to some degree, forming a continuum of phenotypes represented by disease axes, which are composite measures made up of many contributing variables. Supporting this notion, a continuum model of COPD subtypes represented by disease axes derived from principal component analysis was found to be more stable compared to distinct clusters (9), and continuous COPD subtypes represented by emphysema-and airway-predominant disease axes have been identified via factor analysis (10,11). Multiple approaches to dissect COPD heterogeneity are at least partially captured by dividing individuals with significant airflow obstruction into emphysema-predominant and non-emphysema predominant COPD based on CT densitometric assessment of emphysema (12).

In addition to subtyping based on clinical and imaging variables, the increasing availability of and expertise in high-throughput molecular technologies (“omics”) have enabled the molecular subtyping of complex diseases (13). Omic technologies have been extensively used to identify the molecular drivers of heterogeneity in COPD. Genomics has been utilized to develop polygenic risk scores to risk-stratify COPD patients and characterize COPD subtypes (14). Transcriptomics has been used to identify biomarkers of worsening lung function and for risk stratification (15,16). Proteomics has been leveraged to identify biomarkers for emphysema-and non-emphysema-predominant COPD subtypes (17,18) and to develop proteomic risk scores as proxies for respiratory susceptibility (19).

The integration of multiple types of omic data offers a promising avenue to refine molecular subtypes, since unearthing the cooperation between different omic modalities can further elucidate the disease mechanisms defining each subtype. Multi-omic integration methods can be categorized into early, intermediate, and late integration methods (20). Early integration concatenates omic data matrices into one matrix and performs downstream machine learning tasks on this matrix; late integration deploys machine learning models on each data matrix separately, combining their results *post hoc*, and intermediate integration jointly analyzes different omics by projecting them onto a common latent space. Multi-omics factor analysis (MOFA) (21) is a multi-view dimensionality reduction framework that falls in the intermediate integration category. It has emerged as an effective method to identify the major axes of heterogeneity representing continuous subtypes, as well as the molecular drivers of these axes. MOFA has been used in diverse disease contexts, identifying the molecular axes driving inter-individual variability in malignant pleural mesothelioma (22), chronic lymphocytic leukemia (23), acute and chronic coronary syndromes (24), obesity (25), influenza vaccine response (26), amyotrophic lateral sclerosis (27), dysbiotic lung injury (28), HIV infection (29), and immune system development (30). Despite the established clinical heterogeneity of COPD, multi-omics factor analysis has thus far not been used in a large COPD cohort with the purpose of identifying its major disease axes and their drivers.

In the present work, we integrated genomic, transcriptomic, and proteomic data from the Genetic Epidemiology of COPD (COPDGene) study using MOFA. We identified latent factors representing major axes of heterogeneity in COPD and multi-omic contributors to these axes. We tested the association of the latent factors and their top drivers with COPD-related clinical phenotypes including lung function, chest computed tomography (CT), and complete blood count phenotypes, as well as all-cause mortality. We performed quantitative trait loci (QTL) analyses to characterize the common genetic control of gene expression and protein abundance in a latent factor jointly driven by transcriptomics and proteomics. We developed a genomics-driven latent factor-weighted polygenic risk score and tested it on an independent COPD cohort.

## Results

### Multi-omics factor analysis identifies ten latent factors that represent heterogeneity across individuals with moderate-to-very severe COPD

To identify the major axes of heterogeneity across COPD patients, as well as the molecular contributors to these axes, we used multi-omics factor analysis (MOFA) (21). We applied MOFA to genomic (SNP genotyping), transcriptomic (whole blood RNA-seq), and proteomic (plasma SomaScan) data collected in the Genetic Epidemiology of COPD (COPDGene) study (31) **(Figure 1a) (Methods)**. The analysis included a feature set of 7,278 SNPs, 5,000 transcripts, and 4,979 proteins after quality control, preprocessing, and feature selection, and a cohort of 1,872 study participants with moderate to very severe COPD who had at least two omic types available in the five-year follow up visit (Phase 2) of COPDGene **(Figure 1b) (Methods)**. Study cohort characteristics are shown in **Table 1**. The analysis cohort consisted of 437 African American (23.3%) and 1,435 (76.7%) non-Hispanic white individuals who were either former (65.8%) or current (34.2%) smokers as of their Phase 2 visit. Participants showed a balanced representation in terms of sex, self-reported race, and omic data availability across Global Initiative for Chronic Obstructive Lung Disease (GOLD) stages 2 (moderate), 3 (severe), and 4 (very severe), whereas age, body mass index (BMI), smoking status, and pack-years of smoking at Phase 2 were associated with GOLD stage.

**Figure 1:**
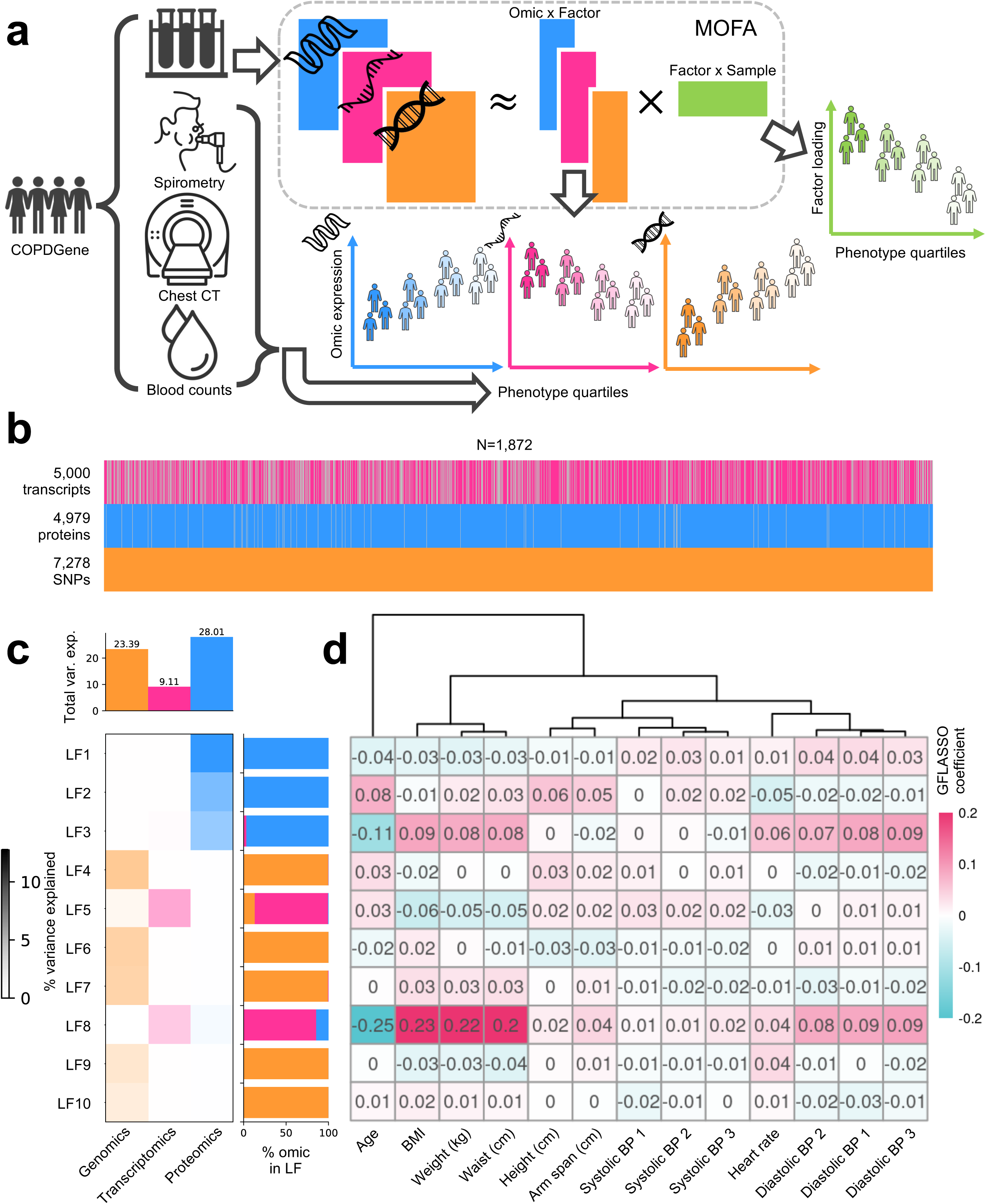
MOFA analysis overview and results. **(a)** Study overview. Orange, pink and blue colors denote genomic, transcriptomic, and proteomic data, respectively. **(b)** Data availability for each individual and feature sets for each omic modality. Each vertical line denotes an individual. Missing data are denoted by grey lines. **(c)** Percent variance explained by latent factor and omic type. Bar plots on top show the total variance explained per omic type. Bar plots on the right show the relative proportion of variance explained by omic type for each latent factor. **(d)** Hierarchically clustered GFLASSO regression coefficients for anthropometric and physiological measurements at Phase 2 visit. Appended numbers indicate separate blood pressure measurements.

**Table 1:** Characteristics of the COPDGene cohort used in the study.

|  | **GOLD Stage** | **2** | **3** | **4** | **Overall** | **p-value** |
| --- | --- | --- | --- | --- | --- | --- |
| **N** |  | 1070 | 567 | 235 | 1872 |  |
| **Genotype available (%)** | Yes | 1070<br>(100.0) | 567<br>(100.0) | 235<br>(100.0) | 1872<br>(100.0) | NA |
| **RNAseq available (%)** | No | 363 (33.9) | 208 (36.7) | 86 (36.6) | 657 (35.1) | 0.471 |
|  | Yes | 707 (66.1) | 359 (63.3) | 149 (63.4) | 1215 (64.9) |  |
| **Proteomics available (%)** | No | 40 (3.7) | 15 (2.6) | 8 (3.4) | 63 (3.4) | 0.506 |
|  | Yes | 1030 (96.3) | 552 (97.4) | 227 (96.6) | 1809 (96.6) |  |
| **Sex (%)** | Male | 568 (53.1) | 318 (56.1) | 141 (60.0) | 1027 (54.9) | 0.122 |
|  | Female | 502 (46.9) | 249 (43.9) | 94 (40.0) | 845 (45.1) |  |
| **Self-reported race (%)** | Non-Hispanic white | 814 (76.1) | 436 (76.9) | 185 (78.7) | 1435 (76.7) | 0.677 |
|  | African American | 256 (23.9) | 131 (23.1) | 50 (21.3) | 437 (23.3) |  |
| **Age at Phase 2 visit (mean (SD))** |  | 67.56<br>(8.41) | 68.77<br>(8.01) | 67.07<br>(7.60) | 67.87<br>(8.21) | 0.005 |
| **BMI at Phase 2 visit (mean (SD))** |  | 28.87<br>(6.28) | 28.00<br>(6.39) | 25.84<br>(6.11) | 28.23<br>(6.36) | <0.001 |
| **Smoking status at Phase 2 visit (%)** | Former smoker | 641 (59.9) | 398 (70.2) | 193 (82.1) | 1232 (65.8) | <0.001 |
|  | Current smoker | 429 (40.1) | 169 (29.8) | 42 (17.9) | 640 (34.2) |  |
| **Pack-Years at Phase 2 visit (mean (SD))** |  | 50.02<br>(23.44) | 55.28<br>(26.71) | 53.24<br>(25.70) | 52.02<br>(24.86) | <0.001 |

MOFA inferred 10 latent factors (LFs), each of which explained at least 2% of the variance in at least one type of omic modality **(Methods) (Table S1, Figure S1a)**. LF1, LF2, and LF3 mainly explained the variance in proteomics, LF4, LF6, LF7, LF9, and LF10 mainly explained the variance in genomics, and LF5 and LF8 mainly explained the variance in transcriptomics **(Figure 1c)**. LF5 and LF8 also explained a secondary omic (>0.5% explained variance) in addition to their primary omic type. Overall, the 10 factors explained 23.4%, 9.1% and 28.0% of the variance in genomic, transcriptomic and proteomic data, respectively **(Figure 1c)**. The identified factors were orthogonal to each other with minimal pairwise correlations **(Figure S1b)**. Graph-guided fused least absolute shrinkage and selection operator (GFLASSO) regression revealed that the identified factors were associated with several correlated anthropometric and physiological measures **(Methods)**. For example, LF3 and LF8 were positively correlated with BMI and blood pressure and negatively correlated with age, and LF2 was positively correlated with age **(Figure 1d)**.

### Latent factors identified by MOFA are associated with COPD-related phenotypes and mortality

To understand the relevance of the latent factors identified by MOFA to COPD-related phenotypes, we tested their association with lung function, chest CT imaging, and blood count phenotypes, as well as all-cause mortality **(Methods)**. For a minimally biased selection of representative phenotypes among multiple correlated phenotypes recorded in COPDGene, we used GFLASSO regression **(Figures S2-S4)**. Based on the hierarchical clustering of GFLASSO coefficients, we chose 13 phenotypes to test their associations with the latent factors **(Methods)**. Five factors were associated (FDR < 0.1) with at least one of the tested phenotypes: LF1, LF3, LF5 and LF8 were associated with blood count phenotypes such as neutrophil counts and hemoglobin levels, LF5, LF8 and LF9 were associated with chest CT imaging phenotypes such as percent emphysema and adjusted lung density, and LF9 was associated with percent-predicted forced expiratory volume in one second (FEV1) **(Figure 2a)**. When we divided individuals into four groups based on the quartiles of each of these phenotypes, the corresponding factor values of these groups were significantly different from each other (adj. ANCOVA p-value < 0.05) **(Figure 2b and Table S2)**. LF3, a proteomics-driven factor associated with neutrophil counts, hemoglobin levels, and six-minute walk distance, also had a modest but significant effect on survival (HR=0.88 [0.80-0.96]), where lower values of LF3 were associated with worse outcomes **(Figure 2c)**. The survival probability of groups of individuals based on their LF3 quartiles differed significantly (log-rank test p-value = 3e-06) (Figure 2d).

**Figure 2:**
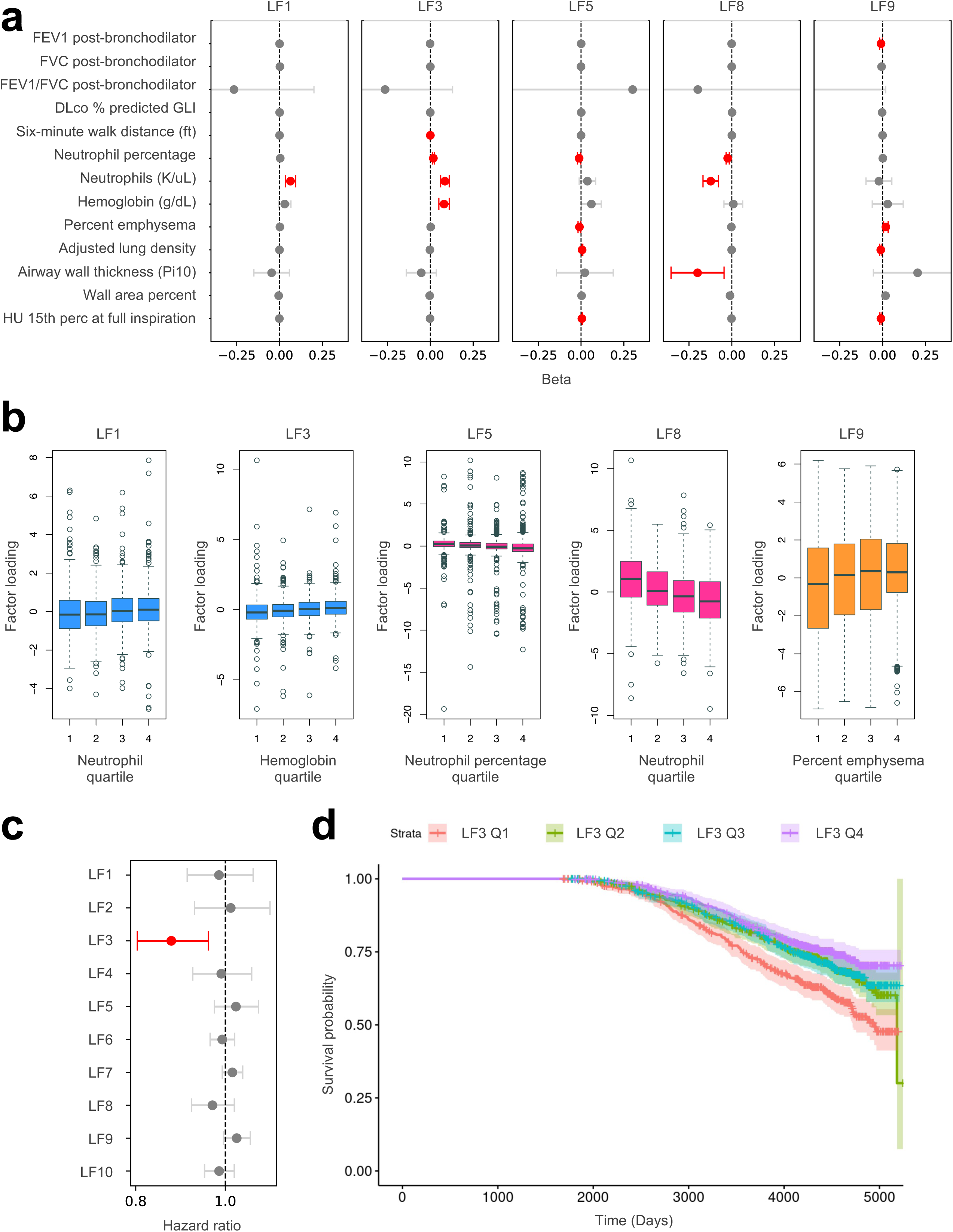
Associations of latent factors (LFs) with COPD-related phenotypes and all-cause mortality. **(a)** Forest plots showing the regression coefficients between LFs and COPD-related phenotypes. Error bars indicate 95% confidence intervals. Associations with a false discovery rate (FDR) less than 0.1 are shown in red. **(b)** Boxplots showing factor loadings versus COPD-related phenotype quartiles. **(c)** Forest plot showing the results of Cox proportional hazards regressions. **(d)** Kaplan-Meier curves showing survival probabilities of groups of individuals based on LF3 quartiles.

### Top omic contributors to latent factors are associated with COPD-related phenotypes and distinguish between patient subgroups

Next, we sought to obtain molecular insights from latent factors by examining their top omic contributors, i.e., the SNPs, transcripts, and proteins with the highest factor loadings. Our hypothesis was that the allele dosages, expression levels, and abundance levels of the top SNPs, transcripts, and proteins for each latent factor would associate with target phenotypes of that factor (i.e., the phenotypes with which the values of a given factor were significantly associated; see **Figure 2a**). To test this hypothesis, we measured the association of the top 100 contributors to each factor with that factor’s target phenotypes **(Methods)**. In 69% of the associated factor-phenotype pairs, more than 70 of the top 100 molecules had significant associations (FDR < 0.1) with the target phenotypes **(Table S3)**. 99 and 98 out of the top 100 proteins of LF1 and LF3, factors that primarily explain the variance in proteomic data, were associated with neutrophils **(Figure S5a)** and hemoglobin **(Figure 3a)**, respectively. For LF5, a factor that primarily explains the variance in RNA-seq and secondarily in genomic data, 94 out of the top 100 genes were significantly associated (FDR < 0.1) with neutrophil percentage **(Figure 3c)**, whereas the associations of top genes and SNPs with chest imaging phenotypes such as percent emphysema and adjusted lung density were weaker with only up to 30 of the top 100 genes and SNPs achieving nominal significance (unadj. p < 0.05) **(Table S3)**. All of the top 100 SNPs of LF9, a factor that primarily explains the variance in genomic data, were located in chromosome 10 downstream of the leucine rich melanocyte differentiation associated (*LRMDA*) gene and the human RNA gene *ENSG00000269256*, and were associated with percent emphysema **(Figure 3e)** and adjusted lung density **(Figure S5c, Table S3),** whereas the associations of top SNPs with FEV1 were weaker with 68 of the top 100 SNPs achieving nominal significance (unadj. p < 0.05) **(Table S3)**.

**Figure 3:**
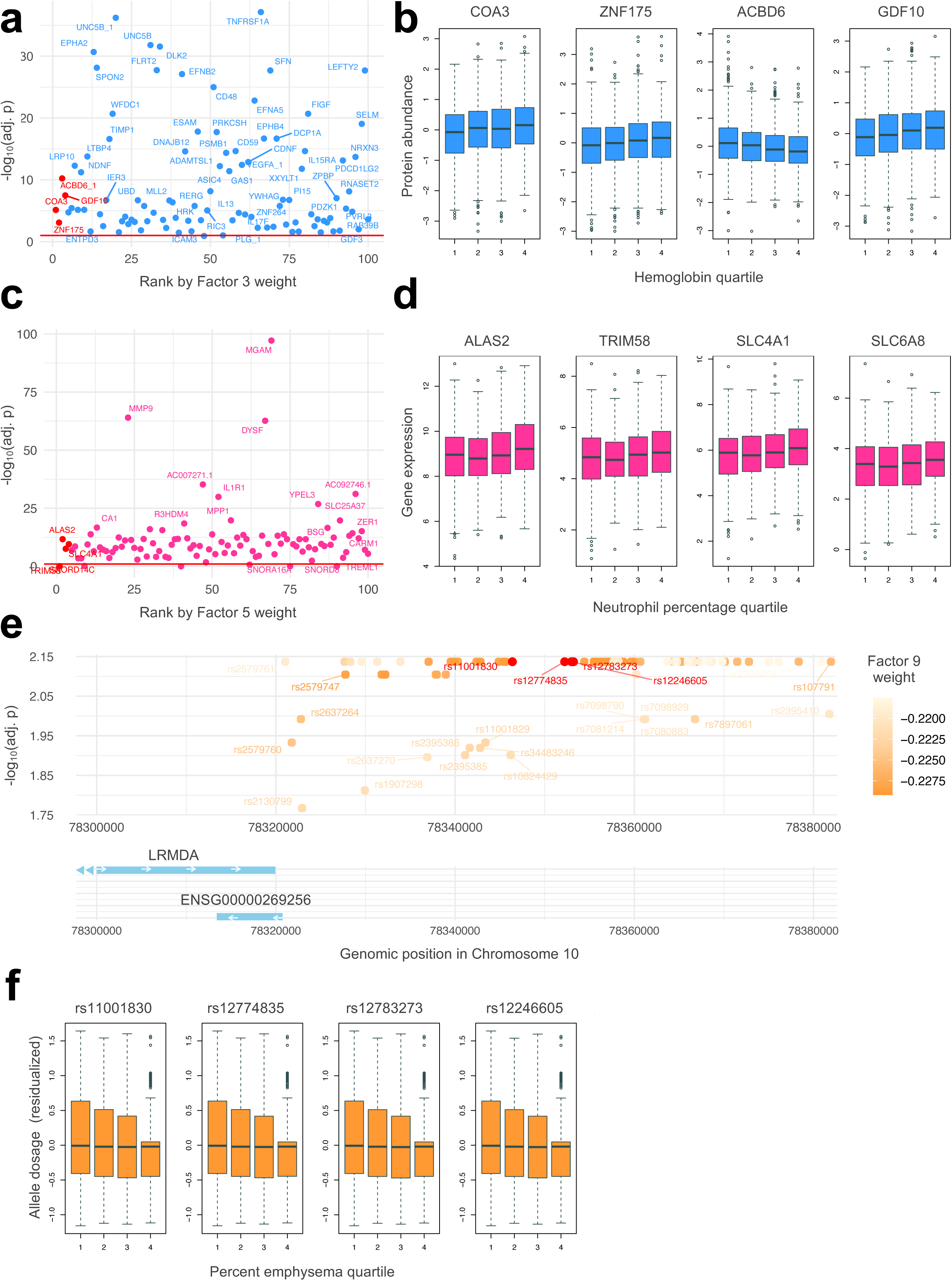
Top omic contributors to latent factors (LFs) and their associations with COPD-related phenotypes. **(a)** Scatter plots showing the top 100 proteins ranked by their LF3 weights versus their respective adjusted p-values of association with hemoglobin levels. The proteins shown in panel (b) are colored in red. **(b)** Boxplots showing the abundance levels of the top four proteins of LF3 with significant associations with hemoglobin levels in individuals grouped according to their hemoglobin quartiles. **(c)** Scatter plots showing the top 100 genes ranked by their LF5 weights versus their respective adjusted p-values of association with neutrophil percentage levels. The genes shown in panel (d) are colored in red. **(d)** Boxplots showing the expression levels of the top four genes of LF5 with significant associations with neutrophil percentage levels in individuals grouped according to their neutrophil percentage quartiles. **(e)** Genomic locations of the top 100 SNPs of LF9 versus their respective adjusted p-values of association with percent emphysema. Darker marker colors indicate higher absolute LF9 weights. The SNPs shown in panel (f) are colored in red. **(f)** Boxplots showing the residualized allele dosages of the top four SNPs of LF9 with significant associations with percent emphysema in individuals grouped according to their percent emphysema quartiles. In panels (a) and (c), the red line indicates an FDR threshold of 0.1.

Similar to factor-phenotype associations, the top omic contributors identified for each factor helped distinguish between groups of individuals based on their phenotype quartiles **(Table S3)**. The abundance levels of the four proteins with the highest factor loadings in LF1, encoded by the genes *CDC42*, *RAB1B*, *RAC1*, and *ERP29*, differentiated significantly (adj. ANCOVA p-value < 0.05) between groups of participants based on their neutrophil quartiles **(Figure S5b).** The abundance levels of the four proteins with the highest factor loadings in LF3, encoded by the genes *COA3*, *ZNF175*, *ACBD6*, and *GDF10*, differentiated significantly (adj. ANCOVA p-value < 0.05) between groups of patients based on their hemoglobin quartiles **(Figure 3b).** The expression levels of four of the five genes with the highest factor loadings in LF5, *ALAS2*, *TRIM58*, *SLC4A1*, and *SLC6A8*, differentiated significantly (adj. ANCOVA p-value < 0.05) between groups of patients based on their neutrophil percentage quartiles **(Figure 3d)**. Finally, the residual allele dosages of the top four SNPs with the highest factor loadings in LF9, rs11001830, rs12774835, rs12783273, and rs12246605, differentiated significantly (adj. ANCOVA p-value < 0.05) between groups of patients based on their percent emphysema and lung density quartiles **(Figure 3f and Table S3)**. Four of the top 100 SNPs of LF9 (rs17384253, 11001829, rs34483246 and rs34763796) were located in an enhancer site (GH10J076582) that was predicted by GeneHancer (32) to interact with the promoter region of *ENSG00000269256* **(Figure S6a)**.

### Integrative genomics of latent factor 8 reveals potential common genetic control of gene expression and protein abundance

LF8 is a latent factor that explained the variance in two omic types, primarily in RNA-seq and secondarily in proteomics. The sets of top 100 gene and protein drivers of LF8 were mutually exclusive **(Table S3)**. Moreover, network proximity analysis **(Methods)** showed that the top genes and proteins contributing to LF8 were not statistically significantly close to one other in the protein-protein interaction (PPI) network **(Table S4)**, suggesting their disparate biological functions (33). The associations of the top genes and proteins with COPD-related outcomes were in line with this observation: While both the top genes and proteins of LF8 had a strong signal for association with neutrophil percentage, a higher portion of the top 100 genes achieved significance (FDR < 0.1) compared to the top 100 proteins (99% vs 64%) **(Figures S5d-e)**. On the other hand, the association with airway wall thickness was driven by proteomics with 37% of top proteins being significant (FDR < 0.1), while only 6% of the top genes achieved nominal significance (unadj. p < 0.05) **(Figures S5f-g)**.

Due to this apparent orthogonality of the transcriptomic and proteomic component of LF8, we investigated whether there could be common genetic control of the non-overlapping top gene and protein contributors to LF8. To assess this, we identified the cis- and trans-expression and protein quantitative trait loci (eQTLs and pQTLs) between all gene-SNP and protein-SNP pairs included in the analysis **(Methods)**. Intersecting the eGenes and pGenes of the identified eQTLs and pQTLs with the genes and proteins in the 90^th^ percentile with respect to their LF8 loadings resulted in 23 genes in 663 eQTLs and 10 proteins in 865 pQTLs with 603 SNPs in common **(Figure 4a)**, focusing on a hotspot in chromosome 6 **(Figure 4b)**. Most of these SNPs mediated in *cis* between the pGene *BTN3A3* and the eGenes *C4A, TRIM10*, and *H3C10*, and in *trans* between *C4A/BTN3A3* and the pGenes *MDK* and *KIR2DL3*, and the eGene *MIR3174* **(Figure 4c, Figure S6b)**. The majority of the remaining trans-QTLs centered around the SNP rs9380069. The gene expression and protein abundance levels were weakly but significantly correlated for two gene-protein pairs that were hubs in the same network community **(Figure 4c inset, see Methods),** namely *C4A* and *BTN3A3* (Pearson’s r = 0.09, p-value = 0.0028) and *TRIM10* and *MDK* (Pearson’s r = −0.08, p-value = 0.0085) **(Figure 4d-e)**.

**Figure 4:**
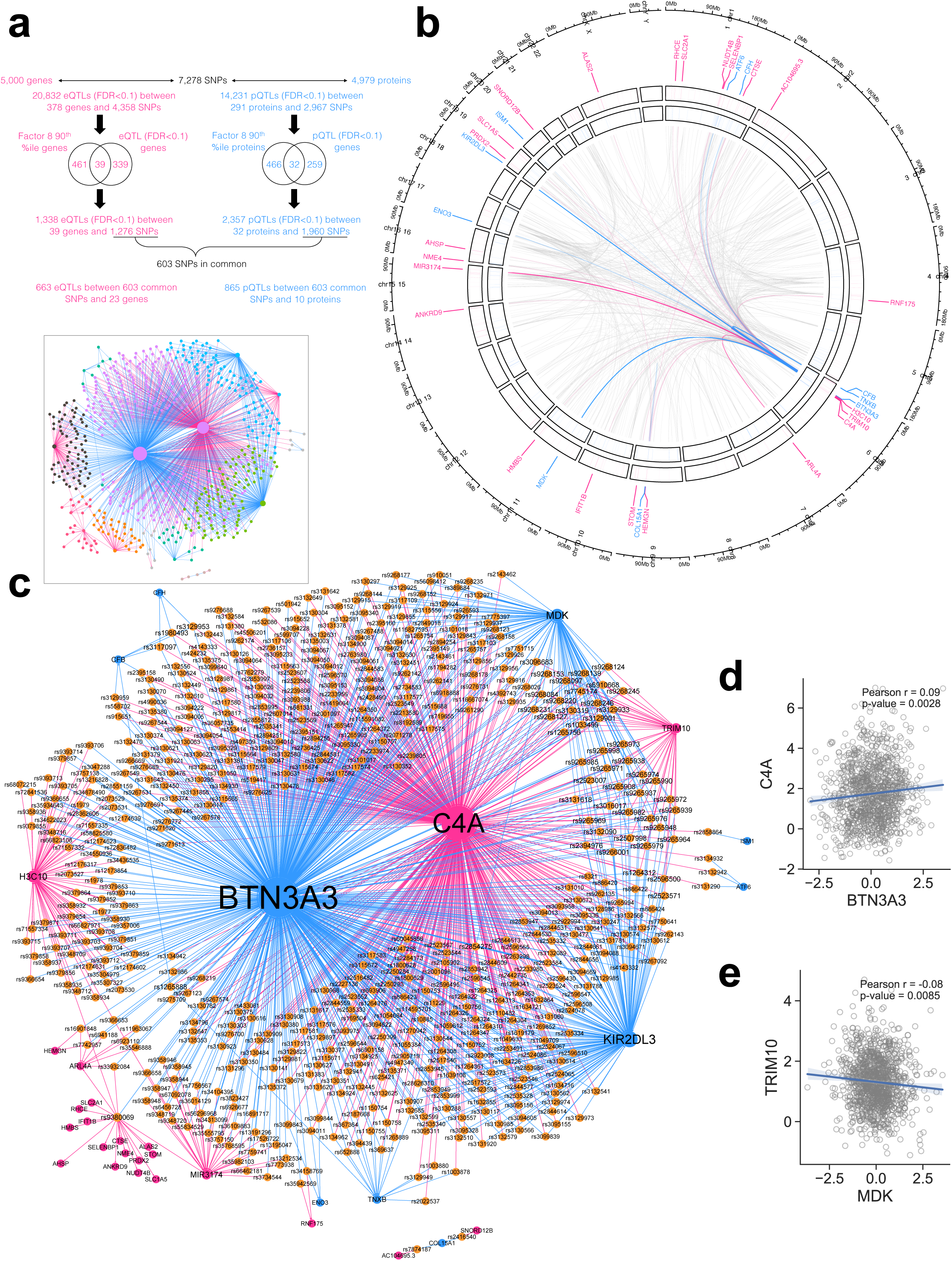
Integrative omics of latent factor 8. **(a)** Summary of quantitative trait loci analyses. eQTL and pQTL denote expression and protein quantitative trait loci, respectively. (b) Circos plot showing the cis- and trans-QTLs. The outer and inner tracks show LF8 weights of the eGenes and pGenes, respectively, with a higher color intensity indicating a higher weight. eQTLs and pQTLs with eGenes and pGenes in the 90^th^ percentile with respect to their LF8 weights are shown as pink and blue edges, respectively, whereas the remaining eQTLs and pQTLs are shown as grey edges. **(c)** QTL network showing the 603 common SNPs connecting the 23 genes and 10 proteins identified. SNPs, genes and proteins are represented by orange, pink, and blue nodes. Pink and blue edges indicate eQTLs and pQTLs. The inset shows the same network with the nodes colored according to their network communities identified by modularity maximization. **(d)** Scatter plot showing the gene expression of C4A against the protein abundance of BTN3A3. Each dot represents an individual in the COPDGene cohort. The regression line is surrounded by the 95% confidence interval. **(e)** Scatter plot showing the gene expression of TRIM10 against the protein abundance of MDK. Each dot represents an individual in the COPDGene cohort. The regression line is surrounded by the 95% confidence interval.

### Polygenic risk scores derived from a genomics-driven latent factor are associated with chest imaging and lung function phenotypes in two independent COPD cohorts

LF9 is the only genomics-driven latent factor that showed significant associations with COPD-related outcomes, and its top SNPs had small and similar effect sizes for these outcomes **(Table S3)**. To aggregate the modest effect sizes of SNPs while accounting for linkage disequilibrium, we devised a polygenic risk score (PRS) specific to the disease axis represented by LF9 **(Methods)**. The LF9-specific PRS values were associated with percent emphysema, wall area, the square root of wall area of a 10-mm luminal perimeter airway (Pi10), and FEV1 in the entire COPDGene cohort for both non-Hispanic white and African American individuals **(Figure 5a)**. To assess the generalizability of the PRS values trained on COPDGene, we sought replication in the Evaluation of COPD Longitudinally to Identify Predictive Surrogate Endpoints (ECLIPSE) study **(Methods)**. Of the COPD-related outcomes that showed associations with the LF9-specific PRS, wall area percentage, percent emphysema, and FEV1 phenotypes were replicated in the ECLIPSE cohort **(Figure 5b)**.

**Figure 5:**
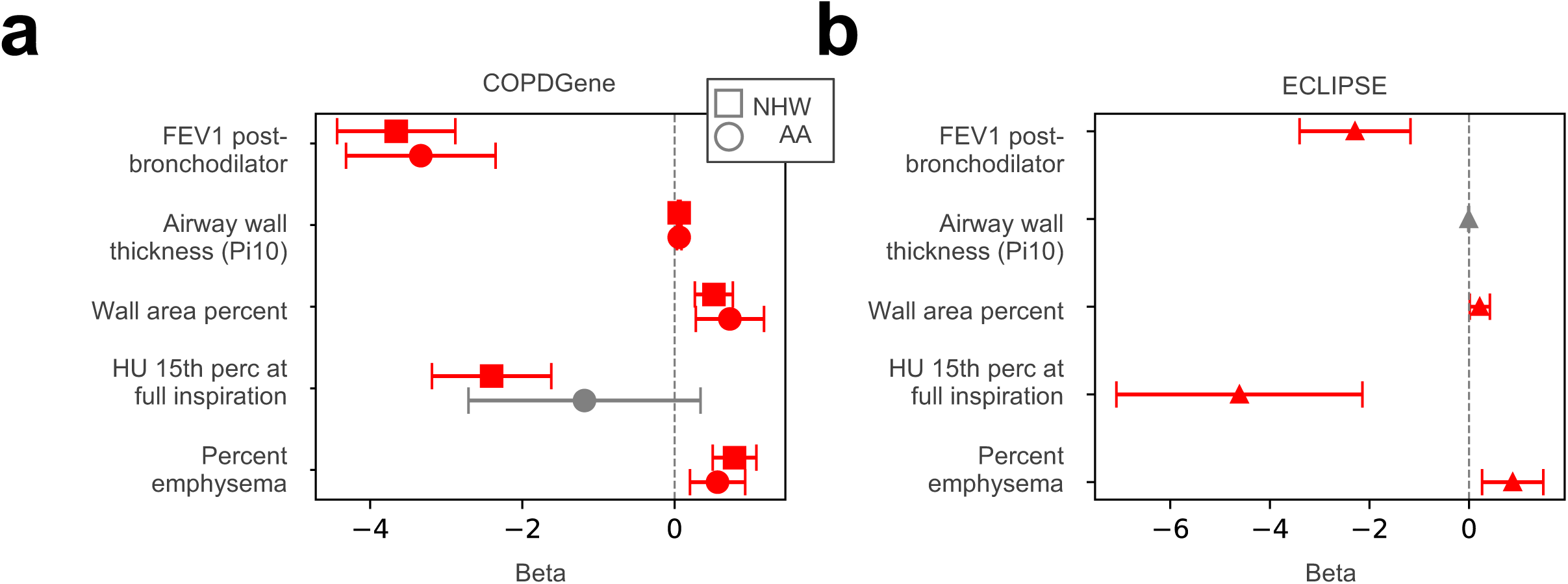
Polygenic risk scores developed using latent factor 9 and their associations with COPD-related outcomes. **(a)** Forest plots showing the regression coefficients between LF9-weighted polygenic risk scores and COPD-related phenotypes in COPDGene. Squares and circles denote non-Hispanic white and African American participants, respectively. **(b)** Forest plots showing the regression coefficients between LF9-weighted polygenic risk scores and COPD-related phenotypes in ECLIPSE. In (a) and (b), error bars indicate 95% confidence intervals. Significant associations are shown in red.

## Discussion

Projecting multi-omic data onto a common latent space offers a feasible means to identify the joint sources of variation between omic types. In this study, we used MOFA to learn the latent factors that represent molecular disease axes for COPD. Five of these latent factors were associated with COPD-related clinical phenotypes, with proteomics-driven factors reflecting blood count phenotypes, transcriptomics-driven factors reflecting CT imaging and blood count phenotypes, and genomics-driven factors reflecting CT imaging and spirometry phenotypes. Latent factor 3, a proteomics-driven factor that is also associated with six-minute walk distance and hemoglobin levels, was associated with all-cause mortality. This finding is consistent with prior literature showing that lower functional status as measured by lower six-minute walk distance is the strongest predictor of all-cause mortality (34), and could potentially indicate low hemoglobin (anemia) or high hemoglobin (erythrocytosis in response to hypoxia). Dividing subjects into quartiles based on these phenotypes further revealed the distinguishing power of these latent factors for continuous subtyping. Together, these results constitute the first step in dissecting the contribution of each omic type to the lung function, chest CT imaging and blood count phenotype axes.

The weight matrices output by MOFA provide valuable insights into which omic elements contribute the most to each disease axis. The top SNPs, transcripts and proteins of a given latent factor ranked by their weight were, in many cases, individually associated with the COPD-related phenotypes that the factor was associated with, suggesting the potential of these top drivers as multi-omic markers of systemic inflammation-, emphysema- and airway-related COPD axes. In line with recent findings from network-based multi-omic analyses on the potential role of inflammation in COPD pathogenesis (35), several inflammatory molecules were identified as top drivers of latent factors. For example, the ratio of the serum levels of tissue inhibitor of metalloproteinase-1 (*TIMP1*) and matrix metalloproteinase-9 (*MMP9*), which are top protein and gene drivers of LF3 and LF5 with strong associations with neutrophils, has recently been proposed as a biomarker for COPD severity (36). As a potential signal for airway-predominant COPD, carcinoembryonic antigen cell adhesion molecule 8 (also known as CD66b), encoded by *CEACAM8*, a top protein driver of LF8 with the strongest association with the square root of wall area of a 10-mm luminal perimeter airway (Pi10), was proposed as a protein biomarker for COPD severity (37) and extracellular vesicle-induced emphysema (38). rs2579760 and rs2579761, which are among the top SNPs of LF9 with significant associations with percent emphysema, are less than 5kb downstream of and in high linkage disequilibrium (D’>0.87) with rs2579762, a SNP located in *LRMDA* that was associated with emphysema-vs. non-emphysema-predominant COPD (18). Some of our findings point towards the potential overlap of disease axes: *CTNNAL1*, a gene that encodes the catenin alpha like 1 protein, and *AHSP*, a gene that encodes the alpha hemoglobin stabilizing protein, which are top gene drivers of LF8 with the strongest association with neutrophil counts and percentage, are also emphysema markers whose effect on emphysema were shown to be mediated by body mass index (BMI) (39), suggesting the putative role of inflammation in this mediation. A further description of the top LF drivers and literature on their COPD relevance can be found in **Table S5**. We note that for some factor-phenotype associations we identified, such as LF5-CT imaging and LF9-lung function phenotypes, no top SNP or gene was individually associated (FDR<0.1) with the target phenotypes of their respective factors. These LF-phenotype associations might instead be collectively driven by the top SNP and gene drivers.

Gene expression and protein abundance are both often strongly genetically regulated, and studies have found moderate levels of overlap between eQTLs and pQTLs, suggesting both common and distinct genetic regulatory processes between genes and the proteins that they encode (40,41). Despite their genetic component, gene expression and protein abundance typically display low correlation due to complex post-transcriptional modifications dictating protein translation, differing decay rates, and other factors (42). We leveraged MOFA’s ability to identify factors explaining variability in multiple omic modalities, and focused on one such factor, LF8, which is a factor with both transcriptomic and proteomic determinants that was associated with neutrophil levels and airway wall thickness. Supporting the above notions in our data, we observed that while the gene and protein drivers of LF8 were distinct from each other, they were tightly genetically regulated by a common set of variants, which enabled us to identify correlations between potentially related pairs of genes and proteins beyond the top drivers of LF8. For example, while complement protein C4, encoded by *C4A*, has been linked to COPD through its decreased serum levels (43), butyrophilin subfamily 3 member A (*BTN3A3*), which has recently been implicated in a severe form of psoriasis (44), has not so far been associated with COPD. Midkine, encoded by *MDK*, was associated with airway remodeling in COPD (45), and while *TRIM10*, encoding tripartite motif 10, has been shown to be a cis-eQTL for COPD (46), its potential role in the proliferation of airway smooth muscle cells has not been investigated. We note that, although not within the core HLA region, *BTN3A3* and *TRIM10* reside nearby in the extended major histocompatibility complex, warranting cautious interpretation of nearby genomic associations.

Variants in linkage disequilibrium (LD), despite being statistically correlated, often have distinct regulatory consequences (47) and allele-specific effects (48). While accounting for LD is typically a pre-processing step in statistical genomic workflows such as GWAS or QTL analyses, in the integrative omics context, performing this step prior to omics integration can mask the potentially diverse functional consequences of variants in LD across different omic layers. Therefore, to avoid the possibility of discarding variants with omic layer-specific functional information, particularly in a setting like MOFA that looks for joint variation across omics, we adopted an approach where we perform LD pruning *post hoc* within a polygenic risk score framework, focusing on a genetically driven factor that associated with COPD-related phenotypes.

The limitations of peripheral blood-based omics in terms of being indirect surrogates of the lung environment (49) and of bulk omics in terms of having a limited ability to shed light on cellular heterogeneity (50) apply to our study as well. We aimed to alleviate the sensitivity of blood-based biomarkers to biological variables (51) by the careful adjustment of these variables in our models. As efforts continue to collect single-cell resolution multi-omic data from the lung tissue, similar integrative omic analyses on high-resolution, tissue-targeted data will be possible, given the recent development of the single-cell-oriented MOFA+ (52). As an important computational caveat, MOFA is a heuristic method that uses variational Bayesian inference that is not guaranteed to converge to a global optimum. To address this and ensure reproducibility, we performed sensitivity analyses and ran multiple models, each with multiple random restarts.

In summary, we identified multi-omic axes of COPD heterogeneity and the molecular drivers of these axes by conducting an integrative omic analysis of a large and well-phenotyped COPD cohort. The resulting COPD disease axes, represented by the latent factors learned by MOFA, were able to distinguish between patient populations based on COPD-related phenotypes, which could be the first step towards a multi-factorial precision diagnosis of COPD that incorporates both multiple omic modalities and complementary clinical measurements such as spirometry and chest CT. On the other hand, the genetic, transcriptomic and proteomic drivers of these disease axes, whose associations with multiple COPD-related outcomes we presented in this study as early evidence of disease relevance, can also act as a resource of potential biomarker candidates which can be investigated in independent mechanistic studies. Moreover, we demonstrated that the rich output provided by MOFA can be leveraged and combined with network methods to investigate the potential interplay between omic layers, providing a foundation for the downstream application of more rigorous statistical genetics approaches such as causal variant identification via fine mapping, or mediation analysis of LFs between SNPs and clinical phenotypes. Similarly, we showed that weights of genetics-driven factors can be used to inform the development of factor-specific polygenic risk scores that can quantify multiple risk factors. Our proof-of-concept applications that go beyond the standard use of MOFA’s outputs can serve as a blueprint for future studies that will aim to use tissue-specific omics and emerging single-cell and spatial data to shed a stronger light onto the heterogeneity of COPD.

## Methods

### Study Populations

The Genetic Epidemiology of COPD (COPDGene) study (31) (ClinicalTrials.gov ID: NCT00608764) is a non-interventional, multicenter, longitudinal study of over 10,000 non-Hispanic white and African American individuals, aged 45-80 years at enrollment, including current and former smokers with ≥ 10 pack years of smoking history, with and without COPD. Detailed anthropometric data, spirometry, CT imaging, and blood samples were collected. The study was extended into a longitudinal study including 5-year (Phase 2) and 10-year (Phase 3) follow up visits.

The Evaluation of COPD Longitudinally to Identify Predictive Surrogate Endpoints (ECLIPSE) study (53) (ClinicalTrials.gov ID: NCT00292552) is a longitudinal observational study of 2,138 individuals, aged 40-75 years, with ≥ 10 pack-years of smoking history. Baseline questionnaire, spirometry, exercise testing, CT imaging, and blood samples were collected. Individuals meeting criteria for GOLD spirometry grades 2-4 (FEV1/FVC < 0.7 and FEV1 % predicted < 80%) were followed up for 3 years.

The GenKOLS (Genetics of Chronic Obstructive Lung Disease) study, a case–control study conducted in Bergen, Norway, was used for polygenic risk score tuning. Eligible individuals had a smoking history greater than 2.5 pack-years, while those with severe alpha-1 antitrypsin deficiency or other chronic lung diseases were excluded. Ethical approvals were obtained from the Regional Committee for Medical Research Ethics (REK Vest), the Norwegian Data Inspectorate, and the Norwegian Department of Health, and all participants provided written informed consent. Genotyping was conducted on the Illumina HumanHap550 array (Illumina, San Diego, CA). Genotypes were imputed with the Michigan Imputation Server using the Haplotype Reference Consortium reference panel.

### Preparation and processing of genomic data

In COPDGene, single nucleotide polymorphism (SNP) genotyping was performed using the Illumina (San Diego, CA) HumanOmniExpress array. Imputation was performed using the Michigan Imputation Server to the Haplotype Reference Consortium (54) and 1000 Genomes Phase I v3 Cosmopolitan reference panels, for non-Hispanic whites and African Americans, respectively. Sex chromosomes were excluded from the genomic data. Allele dosages were adjusted for population substructure in the COPDGene genomic data by regressing out the first six and five ancestry principal components for African-American and non-Hispanic white participants, respectively (55). The Genome Reference Consortium Human Build 37 (GRCh37) was used for variant mapping. Of the over 10,000 COPDGene participants with genomic data, a subset of 1,872 who also had either transcriptomic or proteomic data available in COPDGene Phase 2 were included in the analysis.

In ECLIPSE, SNP genotyping was performed using the Illumina HumanHap 550 V3 (Illumina, San Diego, CA) array. Subjects and markers with call rates <95% were excluded. Imputation was performed using the Michigan Imputation Server and Haplotype Reference Consortium (54) reference panel.

### Preparation and processing of transcriptomic data

RNA extraction and processing of RNA-seq data were previously described in (15,56). Briefly, whole blood samples from the 5-year follow up (Phase 2) of the COPDGene study were collected into PAXgene blood RNA tubes, and total RNA (14,56) was extracted using the PAXgene blood miRNA Kit. After undergoing quality assurance, samples were globin-reduced and cDNA library preparation was performed. 75 bp reads with a mean of 22 million reads per sample were generated using Illumina HiSeq 2500. Count data were filtered to include transcripts with >1 count per million (CPM) in 99% of samples, which were subsequently normalized by log-CPM transformation using the edgeR R package (57). Counts were adjusted for library depth, and batch effects were removed using the limma removeBatchEffects function (58).

### Preparation and processing of proteomic data

Blood proteome levels from the 5-year follow up (Phase 2) of the COPDGene study were measured using the SomaScan v4.0 (5.0K) platform, which uses aptamers (i.e., SOMAmers) to quantify 4,979 unique SOMAmers for 4,776 unique human proteins. The proteomics data were processed using SomaScan’s data standardization procedure that consists of (i) within plate hybridization to control for variability across array signals, (ii) median signal normalization to control for technical variability of replicates within a run, (iii) plate scaling and calibration of SOMAmers to control for inter-assay variation between analytes and batch differences between plates, and (iv) median normalization to a reference using adaptive normalization by maximum likelihood, applied within dilution group, to quality control replicates and individual samples to remove edge effects and technical variance. More details on SomaScan data processing were described previously in (59). For downstream analyses, proteomic data were inverse-normal transformed and study center was regressed out to address demographic confounding (60).

### Multi-omics Factor Analysis (MOFA) implementation

*Feature selection.* In our feature selection, we aimed to keep the number of features comparable across different layers since omics with a significantly larger number of features tend to be overrepresented in the factors identified by MOFA (21). For genomic data, we used the genome-wide significant SNPs from a COPD case-control study (61). Excluding SNPs observed in less than two individuals resulted in 7,313 SNPs in the AA population and 7,319 SNPs in the NHW population, with 7,278 SNPs in common. For transcriptomic data, we matched Ensembl IDs with their respective gene symbols using GENCODE v37 (62), and extracted the top 5,000 mRNAs with the highest variance to be used as features in MOFA. For proteomics, we used all of 4,979 proteins measured by SomaScan.

*Training MOFA models.* We trained MOFA models using the MOFA2 R package (version 1.0.1). We used default model and training parameters except for the following: starting number of factors (set to 15), variance threshold under which factors are dropped (set to 0.02), maximum number of iterations (set to 5,000), and convergence mode (set to “medium”). We tested 18 MOFA models with model choices based on combinations of (i) AA/NHW individuals analyzed separately vs. together, (ii) cohort selection based on individuals with all three omics vs. at least two omics, (iii) pre-MOFA covariate adjustment (unadjusted vs. adjusted for study center-adjusted only vs. adjusted for all covariates). We identified the final model by considering the following criteria: the number of factors with explained variance greater than 2% (higher number of factors preferred), variance explained in each omic type (no dominating omic types preferred), the number of factors with significant COPD-related outcome associations (representation of all phenotype classes preferred), and the number of factors explaining the variance in more than one omic type (higher number of factors preferred) **(Table S1)**. Finally, we ran 10 initializations of the chosen model with different seeds and used the model with the highest evidence lower bound (ELBO) value **(Figure S1a)** for downstream analyses.

### Identifying statistical associations of latent factors and their top contributors with COPD-related outcomes and mortality

For an unbiased selection of representative COPD-related outcomes for further statistical association testing, we performed graph-guided fused least absolute shrinkage and selection operator (GFLASSO) regression (63), which is a unified regularized linear regression framework that takes into account the correlation graph structure among multiple response variables. We calculated the correlation matrices for the three groups of COPD-related phenotypes, namely, lung function, chest CT imaging, and blood count phenotypes, including for each group all phenotypes that were recorded at Phase 2 of the COPDGene study **(Figures S2-4)**. We ran GFLASSO for each phenotype group using the gflasso R package (v.0.0.0.900), performing 5-fold cross-validation to identify the optimal values of the regularization and fusion parameters that minimize the root mean squared error. We then performed hierarchical clustering on the GFLASSO coefficients and used the resulting dendrograms to identify cut-points that delineate subgroups from which to choose one representative phenotype **(Figures S2-4)**. This procedure resulted in 13 representative phenotypes across the three COPD-related phenotype groups.

To identify the factors and the top omic contributors associated with the selected COPD-related outcomes, we fit generalized linear models using the glm() function in the stats R package (v4.0.3), including age, sex, self-reported race, body mass index (BMI), smoking status (current or former smokers), pack-years, and estimated blood cell type proportions as covariates. For factors and omic elements, we used the following regression models, respectively:

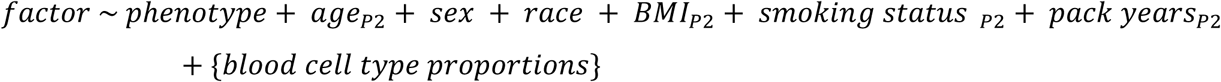

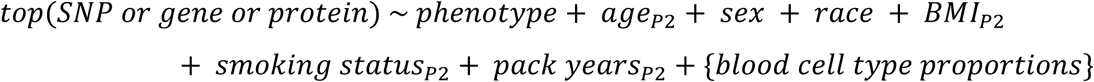

To assess the statistical differences factor values between groups of individuals based on their COPD-related phenotype quantiles, we fit analysis of covariance (ANCOVA) models using the aov() function in R, including the same covariates as above:

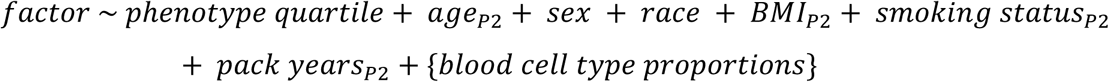

We estimated cell type proportions using the BisqueRNA R package (v.1.0.5) (64) with reference single-cell data from the Human Lung Cell Atlas (https://hlca.ds.czbiohub.org/) (65), and included the estimated proportions for the following cell types as covariates: B, CD4+ Memory/Effector T, CD4+ Naive T, CD8+ Memory/Effector T, CD8+ Naive T, Macrophage, Classical Monocyte, Intermediate Monocyte, Nonclassical Monocyte, Natural Killer, Natural Killer T, Neutrophil, and Proliferating NK/T.

We carried out survival analyses by fitting Cox proportional hazards models using the coxph() function in the survival R package (v.3.2-13), adjusting for the same covariates as above, and plotted Kaplan-Meier curves using the ggsurvplot() function in the survminer R package (v0.4.9). We adjusted all reported p-values for multiple comparisons using the Benjamini-Hochberg procedure and considered as statistically significant a false discovery rate (FDR) of less than 0.1. Unadjudicated all-cause mortality data reported through October 2022 in COPDGene were used for survival analysis.

### Construction of the protein-protein interaction network and network proximity analysis

To quantify the degree of cooperation between the top gene and protein contributors to LF8 on the protein-protein interaction (PPI) network, we used a network-based proximity measure named the closest distance measure (66), defined as

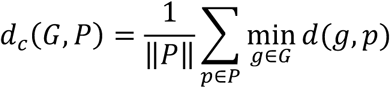

where 𝐺 is the set of top genes contributing to LF8, 𝑃 is the set of top proteins contributing to LF8, and 𝑑(𝑔, 𝑝) is the shortest path length between gene 𝑔 and protein 𝑝. To assess the statistical significance of the measured closest distance between a set of genes and a set of proteins, we performed 200 degree-preserving randomizations of the PPI network and calculated the corresponding 𝑑*_c_*(𝐺, 𝑃) on the randomized networks and calculated empirical p-values, defined as 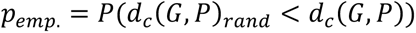. We repeated this procedure for the top 10, 50, 100, and 200 genes and proteins with the highest loadings in LF8. As the PPI network, we used the curated context-independent PPI network provided in (67). After removing self-loops and extracting the largest connected component, the resulting network consisted of 15,865 nodes and 213,714 edges.

### Identifying and visualizing quantitative trait loci in COPDGene

To test the associations between the SNPs, genes, and proteins used in the MOFA analysis, we used the MatrixEQTL R package (v.2.3), including age, sex, self-reported race, body mass index (BMI), smoking status (current or former smokers), pack-years, and the first five ancestry principal components as covariates. We tested all pairwise associations between the 7,278 SNPs and 5,000 genes to identify cis- and trans-eQTLs, and all pairwise associations between the 7,278 SNPs and 4,979 proteins to identify cis- and trans-pQTLs. We adjusted the resulting regression p-values for multiple testing using the Benjamini-Hochberg procedure and retained the eQTLs and pQTLs with FDR < 0.1, which resulted in 20,832 eQTLs between 4,358 SNPs and 378 genes, and 14,231 pQTLs between 291 proteins and 2,967 SNPs. We then intersected these 378 genes and 291 proteins with the top 10% genes and proteins with respect to their LF8 loadings, which resulted in 39 genes and 32 proteins. There were 1,338 eQTLs between these 39 genes and 1,276 SNPs, and 2,357 pQTLs between these 32 proteins and 1,960 SNPs. 603 SNPs were shared between these two sets of loci, which were connected to 23 genes via 663 eQTLs and 10 proteins via 865 pQTLs **(Figure 4a)**. To visualize the genomic positions of the final set of eQTLs and pQTLs, we used the circlize R package (v.0.4.15). To visualize the eQTL and pQTL networks, we used Gephi (v.0.10.1). We used the community detection with modularity optimization feature within Gephi to identify the network-based SNP-gene and SNP-protein clusters.

### Derivation and statistical analysis of the LF9-weighted polygenic risk score

We constructed a polygenic risk score (PRS) using factor weights derived from LF9 and tested its association with the lung function and CT measures associated with LF9. Calculation of a PRS requires a variant, its effect allele, and the corresponding weight. To calculate a LF9-specific PRS, we started with the ∼7,000 variants from a GWAS of COPD (68) that were included in MOFA and refined this set by calculating new weights using lassosum, a penalized regression method that incorporates linkage disequilibrium, estimated from European ancestry participants in the UK Biobank. Hyperparameters (λ and shrinkage) were tuned using the GenKOLS COPD case-control study, selected to avoid parameter training on the COPDGene or ECLIPSE testing cohorts. The shrinkage parameter was derived from the covariance matrix. This procedure accounts for linkage disequilibrium with moderate clumping and thresholding. The finalized PRS was then calculated in COPDGene and ECLIPSE and normalized to have a mean of 0 and standard deviation of 1 for subsequent analyses.

We evaluated the association between the PRS and LF9-associated lung function and CT imaging outcomes, including FEV1 percent predicted, calculated using Global Lung Initiative equations, and quantitative CT measures of emphysema (percent low attenuation area on inspiratory scans (< −950 HU), the 15th percentile of the inspiratory lung density histogram (Perc15)), airway wall thickness (mean wall area percent (WAP), and Pi10 (the square root of wall area for an airway with an internal perimeter of 10 mm)). Linear regression was applied for all outcomes, with adjustment for age, sex, body-mass index, pack-years of smoking, and current smoking status. CT-based outcomes were additionally adjusted for scanner model. Multiple testing was addressed using the Benjamini Hochberg procedure, with a false discovery rate (FDR) threshold of 0.05 for significance.

## Supporting information

Supplemental Figures Combined

## Data Availability

All data produced in the present study are available upon reasonable request to the authors.

## Acknowledgements

We thank Zhonghui Xu and Robert Chase for their help in processing the COPDGene RNA-seq data. AH is supported by NIH grants K25HL150336 and R01HL171141. PJC is supported by NIH grants R01HL124233, R01HL166992, and R01HL171213. DLD is supported by NIH grants K24HL171900, R01HG011393, R01HL178032, and P01HL114501. CPH is supported by NIH grant K24HL173667. KG is supported by NIH grant R01HL155749. EKS is supported by NIH grants P01HL114501, R01HL133135, and R01HL152728. The COPDGene study (NCT00608764) is supported by grants from the NHLBI (U01HL089897 and U01HL089856), by NIH contract 75N92023D00011, and by the COPD Foundation through contributions made to an Industry Advisory Committee that has included AstraZeneca, Bayer Pharmaceuticals, Boehringer-Ingelheim, Genentech, GlaxoSmithKline, Novartis, Pfizer and Sunovion.

## References

1. Soriano JB, Kendrick PJ, Paulson KR, Gupta V, Abrams EM, Adedoyin RA, et al. Prevalence and attributable health burden of chronic respiratory diseases, 1990–2017: a systematic analysis for the Global Burden of Disease Study 2017. Lancet Respir Med [Internet]. 2020 Jun 1 [cited 2025 Sep 13];8(6):585. Available from: https://pmc.ncbi.nlm.nih.gov/articles/PMC7284317/

2. Venkatesan P. GOLD COPD report: 2025 update. Lancet Respir Med [Internet]. 2025 Jan 1 [cited 2025 Nov 19];13(1):e7–8. Available from: https://www.thelancet.com/action/showFullText?pii=S2213260024004132

3. Vestbo J, Hurd SS, Agustí AG, Jones PW, Vogelmeier C, Anzueto A, et al. Global Strategy for the Diagnosis, Management, and Prevention of Chronic Obstructive Pulmonary Disease. 101164/rccm201204-0596PP [Internet]. 2013 Mar 22 [cited 2025 Sep 13];187(4):347–65. Available from: 10.1164/rccm.201204-0596PP?download=true

4. Agusti A, Calverley PMA, Celli B, Coxson HO, Edwards LD, Lomas DA, et al. Characterisation of COPD heterogeneity in the ECLIPSE cohort. Respir Res [Internet]. 2010 Sep 10 [cited 2025 Sep 13];11(1):1–14. Available from: https://respiratory-research.biomedcentral.com/articles/10.1186/1465-9921-11-122

5. Agusti A, Ambrosino N, Blackstock F, Bourbeau J, Casaburi R, Celli B, et al. COPD: Providing the right treatment for the right patient at the right time. Respir Med [Internet]. 2023 Feb 1 [cited 2025 Sep 13];207:107041. Available from: https://www.sciencedirect.com/science/article/pii/S0954611122003067#bib5

6. Barnes PJ, Burney PGJ, Silverman EK, Celli BR, Vestbo J, Wedzicha JA, et al. Chronic obstructive pulmonary disease. Nat Rev Dis Primers [Internet]. 2015 Dec 3 [cited 2025 Sep 13];1(1):1–21. Available from: https://www.nature.com/articles/nrdp201576

7. Lowe KE, Regan EA, Anzueto A, Austin E, Austin JHM, Beaty TH, et al. COPDGene® 2019: Redefining the Diagnosis of Chronic Obstructive Pulmonary Disease. Chronic Obstructive Pulmonary Diseases: Journal of the COPD Foundation [Internet]. 2019 [cited 2025 Sep 13];6(5):384. Available from: https://pmc.ncbi.nlm.nih.gov/articles/PMC7020846/

8. Castaldi PJ, Boueiz A, Yun J, Estepar RSJ, Ross JC, Washko G, et al. Machine Learning Characterization of COPD Subtypes: Insights From the COPDGene Study. Chest [Internet]. 2019 May 1 [cited 2025 Sep 15];157(5):1147. Available from: https://pmc.ncbi.nlm.nih.gov/articles/PMC7242638/

9. Castaldi PJ, Benet M, Petersen H, Rafaels N, Finigan J, Paoletti M, et al. Do COPD subtypes really exist? COPD heterogeneity and clustering in 10 independent cohorts. Thorax [Internet]. 2017 Nov 1 [cited 2025 Nov 19];72(11):998–1006. Available from: https://pubmed.ncbi.nlm.nih.gov/28637835/

10. Kinney GL, Santorico SA, Young KA, Cho MH, Castaldi PJ, San José Estépar R, et al. Identification of Chronic Obstructive Pulmonary Disease Axes That Predict All-Cause Mortality: The COPDGene Study. Am J Epidemiol [Internet]. 2018 Oct 1 [cited 2025 Sep 14];187(10):2109–16. Available from: https://pubmed.ncbi.nlm.nih.gov/29771274/

11. Young KA, Regan EA, Han MLK, Lutz SM, Ragland M, Castaldi PJ, et al. Subtypes of COPD Have Unique Distributions and Differential Risk of Mortality. Chronic Obstructive Pulmonary Diseases: Journal of the COPD Foundation [Internet]. 2019 [cited 2025 Sep 14];6(5):400. Available from: https://pmc.ncbi.nlm.nih.gov/articles/PMC7020845/

12. Castaldi PJ, Xu Z, Young KA, Hokanson JE, Lynch DA, Humphries SM, et al. Heterogeneity and Progression of Chronic Obstructive Pulmonary Disease: Emphysema-Predominant and Non-Emphysema-Predominant Disease. Am J Epidemiol [Internet]. 2023 Oct 1 [cited 2025 Nov 19];192(10):1647–58. Available from: https://pubmed.ncbi.nlm.nih.gov/37160347/

13. Johansson Å, Andreassen OA, Brunak S, Franks PW, Hedman H, Loos RJF, et al. Precision medicine in complex diseases—Molecular subgrouping for improved prediction and treatment stratification. J Intern Med [Internet]. 2023 Oct 1 [cited 2025 Sep 14];294(4):378–96. Available from: /doi/pdf/10.1111/joim.13640

14. Moll M, Sakornsakolpat P, Shrine N, Hobbs BD, DeMeo DL, John C, et al. Chronic obstructive pulmonary disease and related phenotypes: polygenic risk scores in population-based and case-control cohorts. Lancet Respir Med [Internet]. 2020 Jul 1 [cited 2025 Sep 15];8(7):696. Available from: https://pmc.ncbi.nlm.nih.gov/articles/PMC7429152/

15. Moll M, Boueiz A, Ghosh AJ, Saferali A, Lee S, Xu Z, et al. Development of a Blood-based Transcriptional Risk Score for Chronic Obstructive Pulmonary Disease. Am J Respir Crit Care Med [Internet]. 2022 Jan 15 [cited 2025 Aug 12];205(2):161–70. Available from: https://pmc.ncbi.nlm.nih.gov/articles/PMC8787248/

16. Yun JH, Lee S, Srinivasa P, Morrow J, Chase R, Saferali A, et al. An interferon-inducible signature of airway disease from blood gene expression profiling. European Respiratory Journal [Internet]. 2022 May 19 [cited 2025 Aug 13];59(5). Available from: https://publications.ersnet.org/content/erj/59/5/2100569

17. Carolan BJ, Hughes G, Morrow J, Hersh CP, O’Neal WK, Rennard S, et al. The association of plasma biomarkers with computed tomography-assessed emphysema phenotypes. Respir Res [Internet]. 2014 Oct 12 [cited 2025 Sep 15];15(1). Available from: https://pubmed.ncbi.nlm.nih.gov/25306249/

18. Zhang YH, Castaldi PJ, Bowler RP, Pratte KA, Kinney GL, Young KA, et al. Proteomic biomarkers of emphysema-predominant and non-emphysema-predominant chronic obstructive pulmonary disease. EBioMedicine [Internet]. 2025 Jul 1 [cited 2025 Sep 15];117:105800. Available from: https://pmc.ncbi.nlm.nih.gov/articles/PMC12192512/

19. Liu GY, Perry AS, Washko GR, Farber-Eger E, Colangelo LA, Sheng Q, et al. Proteomic Risk Score of Increased Respiratory Susceptibility: A Multicohort Study. 101164/rccm202403-0613OC [Internet]. 2024 Dec 30 [cited 2025 Sep 15];211(1):64–74. Available from: 10.1164/rccm.202403-0613OC?download=true

20. Picard M, Scott-Boyer MP, Bodein A, Périn O, Droit A. Integration strategies of multi-omics data for machine learning analysis. Comput Struct Biotechnol J [Internet]. 2021 Jan 1 [cited 2025 Sep 15];19:3735–46. Available from: https://www.sciencedirect.com/science/article/pii/S2001037021002683#s0075

21. Argelaguet R, Velten B, Arnol D, Dietrich S, Zenz T, Marioni JC, et al. Multi-Omics Factor Analysis—a framework for unsupervised integration of multi-omics data sets. Mol Syst Biol [Internet]. 2018 Jun 20 [cited 2025 Aug 18];14(6):8124. Available from: /doi/pdf/10.15252/msb.20178124?download=true

22. Mangiante L, Alcala N, Sexton-Oates A, Di Genova A, Gonzalez-Perez A, Khandekar A, et al. Multiomic analysis of malignant pleural mesothelioma identifies molecular axes and specialized tumor profiles driving intertumor heterogeneity. Nat Genet [Internet]. 2023 Apr 1 [cited 2025 Sep 15];55(4):607–18. Available from: https://www.nature.com/articles/s41588-023-01321-1

23. Lu J, Cannizzaro E, Meier-Abt F, Scheinost S, Bruch PM, Giles HAR, et al. Multi-omics reveals clinically relevant proliferative drive associated with mTOR-MYC-OXPHOS activity in chronic lymphocytic leukemia. Nat Cancer [Internet]. 2021 Aug 1 [cited 2025 Sep 15];2(8):853. Available from: https://pmc.ncbi.nlm.nih.gov/articles/PMC7611543/

24. Pekayvaz K, Losert C, Knottenberg V, Gold C, van Blokland I V., Oelen R, et al. Multiomic analyses uncover immunological signatures in acute and chronic coronary syndromes. Nat Med [Internet]. 2024 Jun 1 [cited 2025 Sep 15];30(6):1696–710. Available from: https://www.nature.com/articles/s41591-024-02953-4

25. 25. Hinte LC, Castellano-Castillo D, Ghosh A, Melrose K, Gasser E, Noé F, et al. Adipose tissue retains an epigenetic memory of obesity after weight loss. Nature [Internet]. 2024 Dec 12 [cited 2025 Sep 15];636(8042):457. Available from: https://pmc.ncbi.nlm.nih.gov/articles/PMC11634781/

26. Kumar S, Zoodsma M, Nguyen N, Pedroso R, Trittel S, Riese P, et al. Systemic dysregulation and molecular insights into poor influenza vaccine response in the aging population. Sci Adv [Internet]. 2024 Sep 27 [cited 2025 Sep 15];10(39):eadq7006. Available from: https://pmc.ncbi.nlm.nih.gov/articles/PMC11430404/

27. Caldi Gomes L, Hänzelmann S, Hausmann F, Khatri R, Oller S, Parvaz M, et al. Multiomic ALS signatures highlight subclusters and sex differences suggesting the MAPK pathway as therapeutic target. Nat Commun [Internet]. 2024 Dec 1 [cited 2025 Sep 15];15(1):4893. Available from: https://pmc.ncbi.nlm.nih.gov/articles/PMC11161513/

28. Zinter MS, Dvorak CC, Mayday MY, Reyes G, Simon MR, Pearce EM, et al. Pathobiological signatures of dysbiotic lung injury in pediatric patients undergoing stem cell transplantation. Nat Med [Internet]. 2024 Jul 1 [cited 2025 Sep 15];30(7):1982. Available from: https://pmc.ncbi.nlm.nih.gov/articles/PMC11271406/

29. Botey-Bataller J, van Unen N, Blaauw M, Vos WAJW, van Eekeren L, Vadaq N, et al. Genetic and molecular landscape of comorbidities in people living with HIV. Nat Med [Internet]. 2025 Aug 20 [cited 2025 Sep 15];1–10. Available from: https://www.nature.com/articles/s41591-025-03887-1

30. Hill DL, Carr EJ, Rutishauser T, Moncunill G, Campo JJ, Innocentin S, et al. Immune system development varies according to age, location, and anemia in African children. Sci Transl Med [Internet]. 2020 Feb 5 [cited 2025 Sep 15];12(529):eaaw9522. Available from: https://pmc.ncbi.nlm.nih.gov/articles/PMC7738197/

31. Regan EA, Hokanson JE, Murphy JR, Make B, Lynch DA, Beaty TH, et al. Genetic Epidemiology of COPD (COPDGene) Study Design. COPD [Internet]. 2010 [cited 2025 Aug 18];7(1):32. Available from: https://pmc.ncbi.nlm.nih.gov/articles/PMC2924193/

32. Fishilevich S, Nudel R, Rappaport N, Hadar R, Plaschkes I, Stein TI, et al. GeneHancer: genome-wide integration of enhancers and target genes in GeneCards. Database (Oxford) [Internet]. 2017 [cited 2023 Sep 24];2017. Available from: https://pubmed.ncbi.nlm.nih.gov/28605766/

33. Sharan R, Ulitsky I, Shamir R. Network-based prediction of protein function. Mol Syst Biol [Internet]. 2007 Mar 13 [cited 2025 Nov 19];3:1–13. Available from: /doi/pdf/10.1038/msb4100129?download=true

34. Moll M, Qiao D, Regan EA, Hunninghake GM, Make BJ, Tal-Singer R, et al. Machine Learning and Prediction of All-Cause Mortality in COPD. Chest [Internet]. 2020 Sep 1 [cited 2025 Dec 2];158(3):952–64. Available from: https://pubmed.ncbi.nlm.nih.gov/32353417/

35. Sibilio P, Conte F, Huang Y, Castaldi PJ, Hersh CP, DeMeo DL, et al. Correlation-based network integration of lung RNA sequencing and DNA methylation data in chronic obstructive pulmonary disease. Heliyon [Internet]. 2024 May 30 [cited 2025 Nov 24];10(10):e31301. Available from: https://pmc.ncbi.nlm.nih.gov/articles/PMC11130701/

36. Dimic-Janjic S, Hoda MA, Milenkovic B, Kotur-Stevuljevic J, Stjepanovic M, Gompelmann D, et al. The usefulness of MMP-9, TIMP-1 and MMP-9/TIMP-1 ratio for diagnosis and assessment of COPD severity. Eur J Med Res [Internet]. 2023 Dec 1 [cited 2025 Oct 15];28(1):127. Available from: https://pmc.ncbi.nlm.nih.gov/articles/PMC10026402/

37. Soni S, Garner JL, O’Dea KP, Koh M, Finney L, Tirlapur N, et al. Intra-alveolar neutrophil-derived microvesicles are associated with disease severity in COPD. 101152/ajplung000992020 [Internet]. 2021 [cited 2025 Oct 19];320(1):L73–83. Available from: 10.1152/ajplung.00099.2020

38. Margaroli C, Madison MC, Viera L, Russell DW, Gaggar A, Genschmer KR, et al. An in vivo model for extracellular vesicle–induced emphysema. JCI Insight [Internet]. 2022 Feb 22 [cited 2025 Oct 19];7(4). Available from: 10.1172/jci.

39. Suryadevara R, Gregory A, Lu R, Xu Z, Masoomi A, Lutz SM, et al. Blood-based Transcriptomic and Proteomic Biomarkers of Emphysema. 101164/rccm202301-0067OC [Internet]. 2024 Feb 1 [cited 2025 Oct 16];209(3):273–87. Available from: 10.1164/rccm.202301-0067OC?download=true

40. Wu L, Candille SI, Choi Y, Xie D, Jiang L, Li-Pook-Than J, et al. Variation and Genetic Control of Protein Abundance in Humans. Nature [Internet]. 2013 [cited 2025 Oct 20];499(7456):79. Available from: https://pmc.ncbi.nlm.nih.gov/articles/PMC3789121/

41. He B, Shi J, Wang X, Jiang H, Zhu HJ. Genome-wide pQTL analysis of protein expression regulatory networks in the human liver. BMC Biol [Internet]. 2020 Aug 10 [cited 2025 Oct 20];18(1):97. Available from: https://pmc.ncbi.nlm.nih.gov/articles/PMC7418398/

42. Payne SH. The utility of protein and mRNA correlation. Trends Biochem Sci [Internet]. 2014 [cited 2025 Oct 20];40(1):1. Available from: https://pmc.ncbi.nlm.nih.gov/articles/PMC4776753/

43. Westwood JP, Mackay AJ, Donaldson G, Machin SJ, Wedzicha JA, Scully M. The role of complement activation in COPD exacerbation recovery. ERJ Open Res [Internet]. 2016 Oct 19 [cited 2025 Oct 21];2(4). Available from: https://publications.ersnet.org/content/erjor/2/4/00027-2016

44. Lee CC, Huang YH, Chi CC, Chung WH, Chen CB. Generalized pustular psoriasis: immunological mechanisms, genetics, and emerging therapeutics. Trends Immunol [Internet]. 2025 Jan 1 [cited 2025 Oct 21];46(1):74–89. Available from: https://www.cell.com/action/showFullText?pii=S147149062400303X

45. Deng T, Huang Q, Lin K, Qian J, Li Q, Li L, et al. Midkine-Notch2 Pathway Mediates Excessive Proliferation of Airway Smooth Muscle Cells in Chronic Obstructive Lung Disease. Front Pharmacol [Internet]. 2022 Jun 14 [cited 2025 Oct 21];13:794952–794952. Available from: https://europepmc.org/articles/PMC9239375

46. Moll M, Jackson VE, Yu B, Grove ML, London SJ, Gharib SA, et al. A systematic analysis of protein-altering exonic variants in chronic obstructive pulmonary disease. Am J Physiol Lung Cell Mol Physiol [Internet]. 2021 Jul 1 [cited 2025 Oct 21];321(1):L130. Available from: https://pmc.ncbi.nlm.nih.gov/articles/PMC8321852/

47. Grubert F, Zaugg JB, Kasowski M, Ursu O, Spacek D V., Martin AR, et al. Genetic Control of Chromatin States in Humans Involves Local and Distal Chromosomal Interactions. Cell [Internet]. 2015 Aug 27 [cited 2025 Oct 22];162(5):1051. Available from: https://pmc.ncbi.nlm.nih.gov/articles/PMC4556133/

48. Pugalenthi PV, He B, Xie L, Nho K, Saykin AJ, Yan J. Deciphering the tissue-specific functional effect of Alzheimer risk SNPs with deep genome annotation. BioData Min [Internet]. 2024 Dec 1 [cited 2025 Oct 22];17(1):1–13. Available from: https://biodatamining.biomedcentral.com/articles/10.1186/s13040-024-00400-1

49. Balnis J, Lauria EJM, Yucel R, Singer HA, Alisch RS, Jaitovich A. Peripheral Blood Omics and Other Multiplex-based Systems in Pulmonary and Critical Care Medicine. Am J Respir Cell Mol Biol [Internet]. 2023 Oct 1 [cited 2025 Oct 22];69(4):383. Available from: https://pmc.ncbi.nlm.nih.gov/articles/PMC10557924/

50. Lim J, Park C, Kim M, Kim H, Kim J, Lee DS. Advances in single-cell omics and multiomics for high-resolution molecular profiling. Exp Mol Med [Internet]. 2024 Mar 1 [cited 2025 Oct 22];56(3):515–26. Available from: https://www.nature.com/articles/s12276-024-01186-2

51. Regan EA, Hersh CP, Castaldi PJ, DeMeo DL, Silverman EK, Crapo JD, et al. Omics and the Search for Blood Biomarkers in Chronic Obstructive Pulmonary Disease. Insights from COPDGene. Am J Respir Cell Mol Biol [Internet]. 2019 Aug 1 [cited 2025 Oct 22];61(2):143. Available from: https://pmc.ncbi.nlm.nih.gov/articles/PMC6670029/

52. Argelaguet R, Arnol D, Bredikhin D, Deloro Y, Velten B, Marioni JC, et al. MOFA+: A statistical framework for comprehensive integration of multi-modal single-cell data. Genome Biol [Internet]. 2020 May 11 [cited 2025 Oct 22];21(1):1–17. Available from: https://genomebiology.biomedcentral.com/articles/10.1186/s13059-020-02015-1

53. Vestbo J, Anderson W, Coxson HO, Crim C, Dawber F, Edwards L, et al. Evaluation of COPD Longitudinally to Identify Predictive Surrogate End-points (ECLIPSE). European Respiratory Journal [Internet]. 2008 Apr [cited 2025 Aug 29];31(4):869–73. Available from: https://pubmed.ncbi.nlm.nih.gov/18216052/

54. Loh PR, Danecek P, Palamara PF, Fuchsberger C, Reshef YA, Finucane HK, et al. Reference-based phasing using the Haplotype Reference Consortium panel. Nat Genet [Internet]. 2016 Nov 1 [cited 2025 Aug 29];48(11):1443–8. Available from: https://pubmed.ncbi.nlm.nih.gov/27694958/

55. Cho MH, McDonald MLN, Zhou X, Mattheisen M, Castaldi PJ, Hersh CP, et al. Risk loci for chronic obstructive pulmonary disease: A genome-wide association study and meta-analysis. Lancet Respir Med [Internet]. 2014 [cited 2025 Dec 1];2(3):214–25. Available from: https://pubmed.ncbi.nlm.nih.gov/24621683/

56. Ryu MH, Yun JH, Morrow JD, Saferali A, Castaldi P, Chase R, et al. Blood Gene Expression and Immune Cell Subtypes Associated with Chronic Obstructive Pulmonary Disease Exacerbations. Am J Respir Crit Care Med [Internet]. 2023 Aug 1 [cited 2025 Nov 19];208(3):247. Available from: https://pmc.ncbi.nlm.nih.gov/articles/PMC10395718/

57. Robinson MD, McCarthy DJ, Smyth GK. edgeR: a Bioconductor package for differential expression analysis of digital gene expression data. Bioinformatics [Internet]. 2010 Jan 1 [cited 2025 Aug 29];26(1):139–40. Available from: 10.1093/bioinformatics/btp616

58. Ritchie ME, Phipson B, Wu D, Hu Y, Law CW, Shi W, et al. limma powers differential expression analyses for RNA-sequencing and microarray studies. Nucleic Acids Res [Internet]. 2015 Apr 20 [cited 2025 Aug 29];43(7):e47–e47. Available from: 10.1093/nar/gkv007

59. Serban KA, Pratte KA, Strange C, Sandhaus RA, Turner AM, Beiko T, et al. Unique and shared systemic biomarkers for emphysema in Alpha-1 Antitrypsin deficiency and chronic obstructive pulmonary disease. EBioMedicine [Internet]. 2022 Oct 1 [cited 2025 Aug 29];84. Available from: https://pubmed.ncbi.nlm.nih.gov/36155958/

60. Konigsberg IR, Vu T, Liu W, Litkowski EM, Pratte KA, Vargas LB, et al. Proteomic Networks and Related Genetic Variants Associated with Smoking and Chronic Obstructive Pulmonary Disease. medRxiv [Internet]. 2024 Feb 28 [cited 2025 Sep 1];2024.02.26.24303069. Available from: https://pmc.ncbi.nlm.nih.gov/articles/PMC10925350/

61. Sakornsakolpat P, Prokopenko D, Lamontagne M, Reeve NF, Guyatt AL, Jackson VE, et al. Genetic landscape of chronic obstructive pulmonary disease identifies heterogeneous cell-type and phenotype associations. Nat Genet [Internet]. 2019 Mar 25 [cited 2019 Mar 6];51(3):494–505. Available from: http://www.nature.com/articles/s41588-018-0342-2

62. Frankish A, Carbonell-Sala S, Diekhans M, Jungreis I, Loveland JE, Mudge JM, et al. GENCODE: reference annotation for the human and mouse genomes in 2023. Nucleic Acids Res [Internet]. 2023 Jan 6 [cited 2025 Sep 1];51(D1):D942–9. Available from: 10.1093/nar/gkac1071

63. Kim S, Sohn KA, Xing EP. A multivariate regression approach to association analysis of a quantitative trait network. Bioinformatics [Internet]. 2009 Jun 15 [cited 2025 Sep 1];25(12):i204–12. Available from: 10.1093/bioinformatics/btp218

64. Jew B, Alvarez M, Rahmani E, Miao Z, Ko A, Garske KM, et al. Accurate estimation of cell composition in bulk expression through robust integration of single-cell information. Nat Commun [Internet]. 2020 Dec 1 [cited 2025 Sep 1];11(1):1–11. Available from: https://www.nature.com/articles/s41467-020-15816-6

65. Travaglini KJ, Nabhan AN, Penland L, Sinha R, Gillich A, Sit R V., et al. A molecular cell atlas of the human lung from single-cell RNA sequencing. Nature 2020 587:7835 [Internet]. 2020 Nov 18 [cited 2025 Sep 1];587(7835):619–25. Available from: https://www.nature.com/articles/s41586-020-2922-4

66. Guney E, Menche J, Vidal M, Barábasi AL. Network-based in silico drug efficacy screening. Nat Commun. 2016;7:10331.

67. Cheng F, Kovács IA, Barabási AL. Network-based prediction of drug combinations. Nat Commun [Internet]. 2019 Dec 13 [cited 2021 May 16];10(1):1197. Available from: http://www.nature.com/articles/s41467-019-09186-x

68. Moll M, Sakornsakolpat P, Shrine N, Hobbs BD, DeMeo DL, John C, et al. Chronic obstructive pulmonary disease and related phenotypes: polygenic risk scores in population-based and case-control cohorts. Lancet Respir Med [Internet]. 2020 Jul 1 [cited 2025 Sep 3];8(7):696–708. Available from: https://www.thelancet.com/action/showFullText?pii=S2213260020301016

