## Supplemental Figures Combined for "Integrative multi-omic analyses identify major axes of heterogeneity in chronic obstructive pulmonary disease and uncover their molecular contributors"

### Supplementary Figures

**Supplementary Figure and Table Legends**

**Figure S1: (a)** Evidence lower bound (ELBO) value of MOFA runs. Each point indicates an independentinitialization. **(b)** Dot plot showing the correlations between the 10 latent factors identified by MOFA.

**Figure S2: (a)** Dot plot showing the correlations between COPDGene Phase 2 CT imaging measurements. **(b)** Hierarchically clustered GFLASSO regression coefficients for CT imaging measurements at the Phase 2 visit.

**Figure S3: (a)** Dot plot showing the correlations between COPDGene Phase 2 lung function measurements. **(b)** Hierarchically clustered GFLASSO regression coefficients for lung function measurements at the Phase 2 visit.

**Figure S4: (a)** Dot plot showing the correlations between COPDGene Phase 2 complete blood count measurements. **(b)** Hierarchically clustered GFLASSO regression coefficients for complete blood count measurements at the Phase 2 visit.

**Figure S5: (a)** Scatter plots showing the top 100 proteins ranked by their LF1 weights versus their respective adjusted p-values of association with neutrophil counts. The proteins shown in panel (b) are colored in red. **(b)** Boxplots showing the abundance levels of the top four proteins of LF1 with significant associations with neutrophil counts in individuals grouped according to their neutrophil quartiles. **(c)** Genomic locations of the top 100 SNPs in LF9 versus their respective adjusted p-values of association with adjusted lung density. Point colors indicate LF9 weights. The top four SNPs of LF9 with significant associations with adjusted lung density are colored in red. **(d)** Scatter plots showing the top 100 genes ranked by their LF8 weights versus their respective adjusted p-values of association with neutrophil percentage. The top four genes are colored in red. **(e)** Scatter plots showing the top 100 proteins ranked by their LF8 weights versus their respective adjusted p-values of association with neutrophil percentage. The top four proteins are colored in red. **(f)** Scatter plots showing the top 100 genes ranked by their LF8 weights versus their respective adjusted p-values of association with airway wall thickness. The top four genes are colored in red. **(g)** Scatter plots showing the top 100 proteins ranked by their LF8 weights versus their respective adjusted p-values of association with airway wall thickness. The top four proteins are colored in red.

**Figure S6: (a)** UCSC Genome Browser tracks showing chromosomal location, top 100 LF9 SNPs, GeneHancer-predicted promoter-enhancer interactions, and H3K27Ac marks present in the genomic region chr10:78,300,000-78,380,000. Red rectangle denotes the enhancer region overlapping with the top LF9 SNPs. **(b)** QTL network showing the 603 common SNPs connecting the 23 genes and 10 proteins identified. SNPs, genes and proteins are colored according to thee chromosome in which they reside. Pink and blue edges indicate eQTLs and pQTLs. The legend shows the node colors and percentage of nodes corresponding to each chromosome.

**Table S1:** Descriptions and relevant outputs of the 18 MOFA models investigated.

**Table S2:** ANCOVA p-values of each latent factor-COPD phenotype pair when individuals are grouped into quartiles based on these phenotypes.

**Table S3:** Omic-phenotype association p-values of the top 100 SNPs/genes/proteins for each significant (FDR<0.1) latent factor-phenotype association. Betas, p-values, and adjusted p-values (FDR) are obtained from regression models, whereas the reported adjusted and unadjusted ANCOVA p-values are obtained from the LF-phenotype quartile comparisons.

**Table S4:** Network proximity z-scores and empirical p-values for the top 10, 50, 100, and 200 genes and proteins with the highest loadings in LF8.

**Table S5:** Summary of literature and references supporting the potential relationship between top SNPs, genes and proteins of a given latent factor and their associated COPD-related phenotypes.

**a**

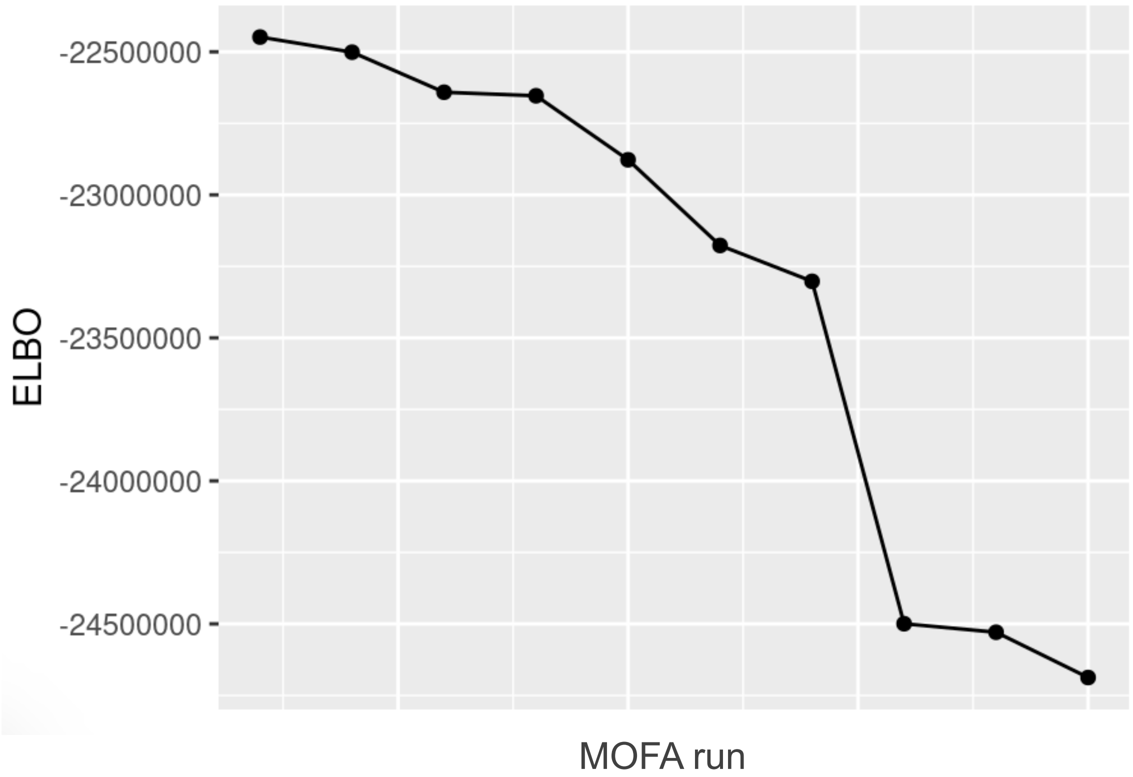

-22500000
-23000000
-23500000
-24000000
-24500000
ELBO
MOFA run


**b**

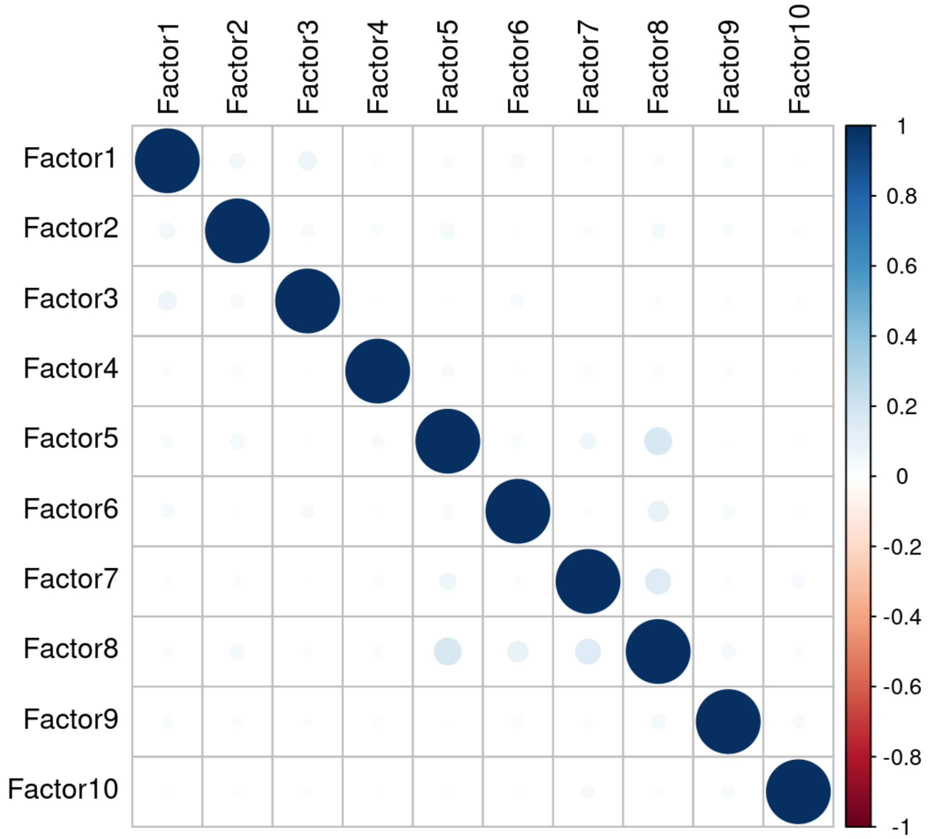

Factor1
Factor2
Factor3
Factor4
Factor5
Factor6
Factor7
Factor8
Factor9
Factor10
1
0.8
0.6
0.4
0.2
0
-0.2
-0.4
-0.6
-0.8
-1


**Figure S1**

a

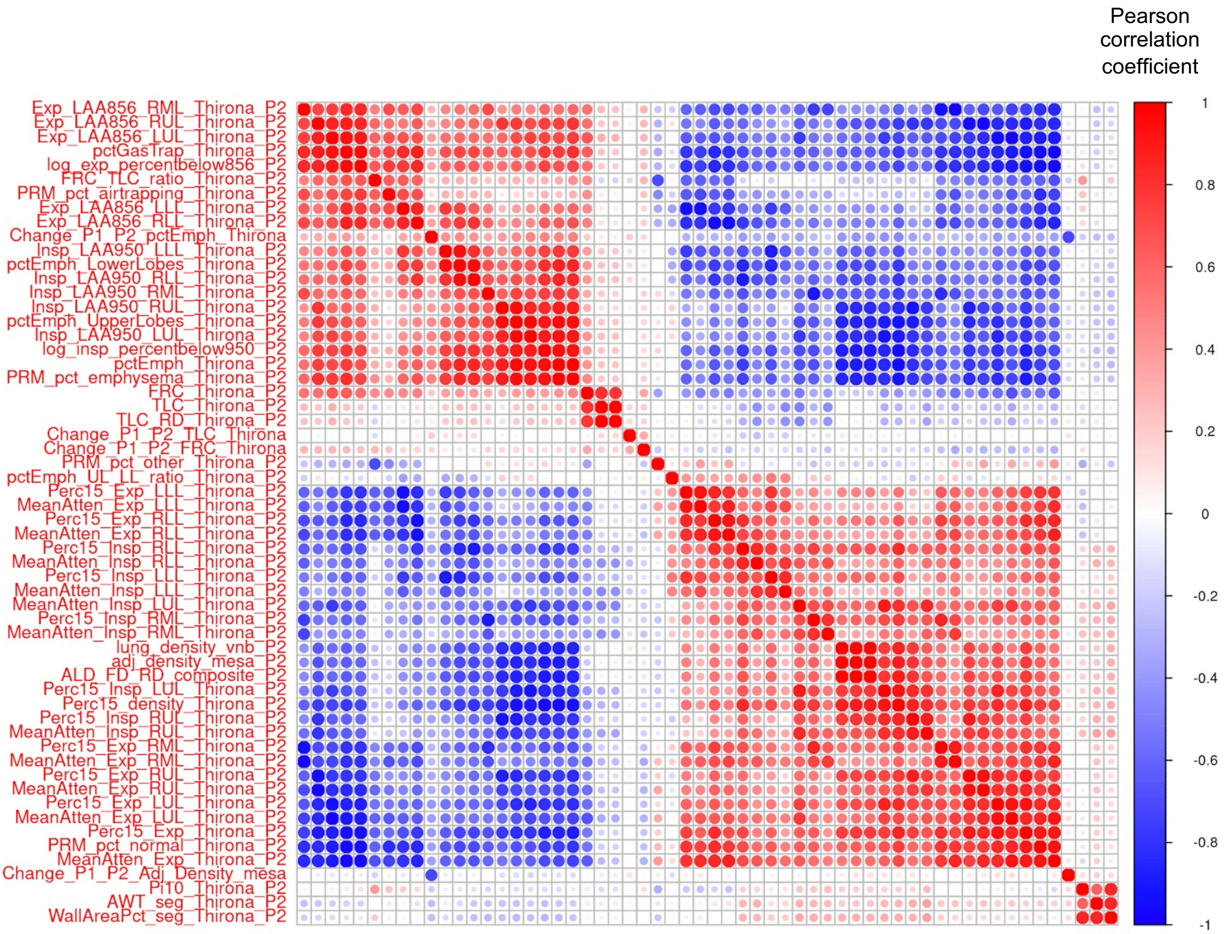
Pearson correlation coefficient
1
0.8
0.6
0.4
0.2
0
-0.2
-0.4
-0.6
-0.8
-1
Exp_LAA856_RML_Thirona_P2
Exp_LAA856_RUL_Thirona_P2
Exp_LAA856_LUL_Thirona_P2
pctGasTrap_Thirona_P2
log_exp_percentbelow856_P2
FRC_TLC_ratio_Thirona_P2
PRM_pct_airtrapping_Thirona_P2
Exp_LAA856_LLL_Thirona_P2
Exp_LAA856_RLL_Thirona_P2
Change_P1_P2_pctEmph_Thirona
Insp_LAA950_LLL_Thirona_P2
pctEmph_LowerLobes_Thirona_P2
Insp_LAA950_RLL_Thirona_P2
Insp_LAA950_RML_Thirona_P2
Insp_LAA950_RUL_Thirona_P2
pctEmph_UpperLobes_Thirona_P2
Insp_LAA950_LUL_Thirona_P2
log_insp_percentbelow950_P2
pctEmph_Thirona_P2
PRM_pct_emphysema_Thirona_P2
FRC_Thirona_P2
TLC_Thirona_P2
TLC_RD_Thirona_P2
Change_P1_P2_TLC_Thirona
Change_P1_P2_FRC_Thirona
PRM_pct_other_Thirona_P2
pctEmph_UL_LL_ratio_Thirona_P2
Perc15_Exp_LLL_Thirona_P2
MeanAtten_Exp_LLL_Thirona_P2
Perc15_Exp_RLL_Thirona_P2
MeanAtten_Exp_RLL_Thirona_P2
Perc15_Insp_RLL_Thirona_P2
MeanAtten_Insp_RLL_Thirona_P2
Perc15_Insp_LLL_Thirona_P2
MeanAtten_Insp_LLL_Thirona_P2
MeanAtten_Insp_LUL_Thirona_P2
Perc15_Insp_RML_Thirona_P2
MeanAtten_Insp_RML_Thirona_P2
lung_density_vnb_P2
adj_density_mesa_P2
ALD_FD_RD_composite_P2
Perc15_Insp_LUL_Thirona_P2
Perc15_density_Thirona_P2
Perc15_Insp_RUL_Thirona_P2
MeanAtten_Insp_RUL_Thirona_P2
Perc15_Exp_RML_Thirona_P2
MeanAtten_Exp_RML_Thirona_P2
Perc15_Exp_RUL_Thirona_P2
MeanAtten_Exp_RUL_Thirona_P2
Perc15_Exp_LUL_Thirona_P2
MeanAtten_Exp_LUL_Thirona_P2
Perc15_Exp_Thirona_P2
PRM_pct_normal_Thirona_P2
MeanAtten_Exp_Thirona_P2
Change_P1_P2_Adj_Density_mesa
Pi10_Thirona_P2
AWT_seg_Thirona_P2
WallAreaPct_seg_Thirona_P2

b

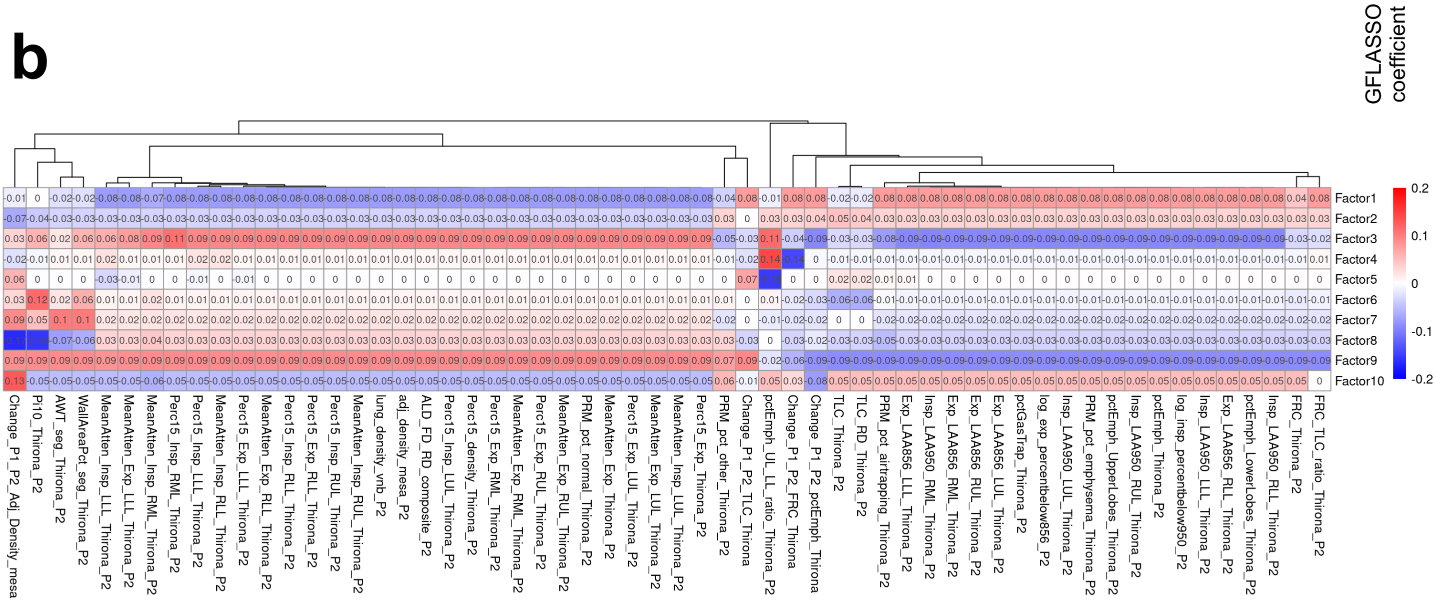
GFLASSO coefficient
0.2
0.1
0
-0.1
-0.2
Factor1
Factor2
Factor3
Factor4
Factor5
Factor6
Factor7
Factor8
Factor9
Factor10
Change_P1_P2_Adj_Density_mesa
Pi10_Thirona_P2
AWT_seg_Thirona_P2
WallAreaPct_seg_Thirona_P2
MeanAtten_Insp_LLL_Thirona_P2
MeanAtten_Exp_LLL_Thirona_P2
MeanAtten_Insp_RML_Thirona_P2
Perc15_Insp_RML_Thirona_P2
Perc15_Insp_LLL_Thirona_P2
MeanAtten_Insp_RLL_Thirona_P2
Perc15_Exp_LLL_Thirona_P2
MeanAtten_Exp_RLL_Thirona_P2
Perc15_Insp_RLL_Thirona_P2
Perc15_Exp_RLL_Thirona_P2
Perc15_Insp_RUL_Thirona_P2
MeanAtten_Insp_RUL_Thirona_P2
lung_density_vnb_P2
adj_density_mesa_P2
ALD_FD_RD_composite_P2
Perc15_Insp_LUL_Thirona_P2
Perc15_density_Thirona_P2
Perc15_Exp_RML_Thirona_P2
MeanAtten_Exp_RML_Thirona_P2
Perc15_Exp_RUL_Thirona_P2
MeanAtten_Exp_RUL_Thirona_P2
PRM_pct_normal_Thirona_P2
MeanAtten_Exp_Thirona_P2
Perc15_Exp_LUL_Thirona_P2
MeanAtten_Exp_LUL_Thirona_P2
MeanAtten_Insp_LUL_Thirona_P2
Perc15_Exp_Thirona_P2
PRM_pct_other_Thirona_P2
Change_P1_P2_TLC_Thirona
pctEmph_UL_LL_ratio_Thirona_P2
Change_P1_P2_FRC_Thirona
Change_P1_P2_pctEmph_Thirona
TLC_Thirona_P2
TLC_RD_Thirona_P2
PRM_pct_airtrapping_Thirona_P2
Exp_LAA856_LLL_Thirona_P2
Insp_LAA950_RML_Thirona_P2
Exp_LAA856_RML_Thirona_P2
Exp_LAA856_RUL_Thirona_P2
Exp_LAA856_LUL_Thirona_P2
pctGasTrap_Thirona_P2
log_exp_percentbelow856_P2
Insp_LAA950_LUL_Thirona_P2
PRM_pct_emphysema_Thirona_P2
pctEmph_UpperLobes_Thirona_P2
Insp_LAA950_RUL_Thirona_P2
pctEmph_Thirona_P2
log_insp_percentbelow950_P2
Insp_LAA950_LLL_Thirona_P2
Exp_LAA856_RLL_Thirona_P2
pctEmph_LowerLobes_Thirona_P2
Insp_LAA950_RLL_Thirona_P2
FRC_Thirona_P2
FRC_TLC_ratio_Thirona_P2

**Figure S2**

a

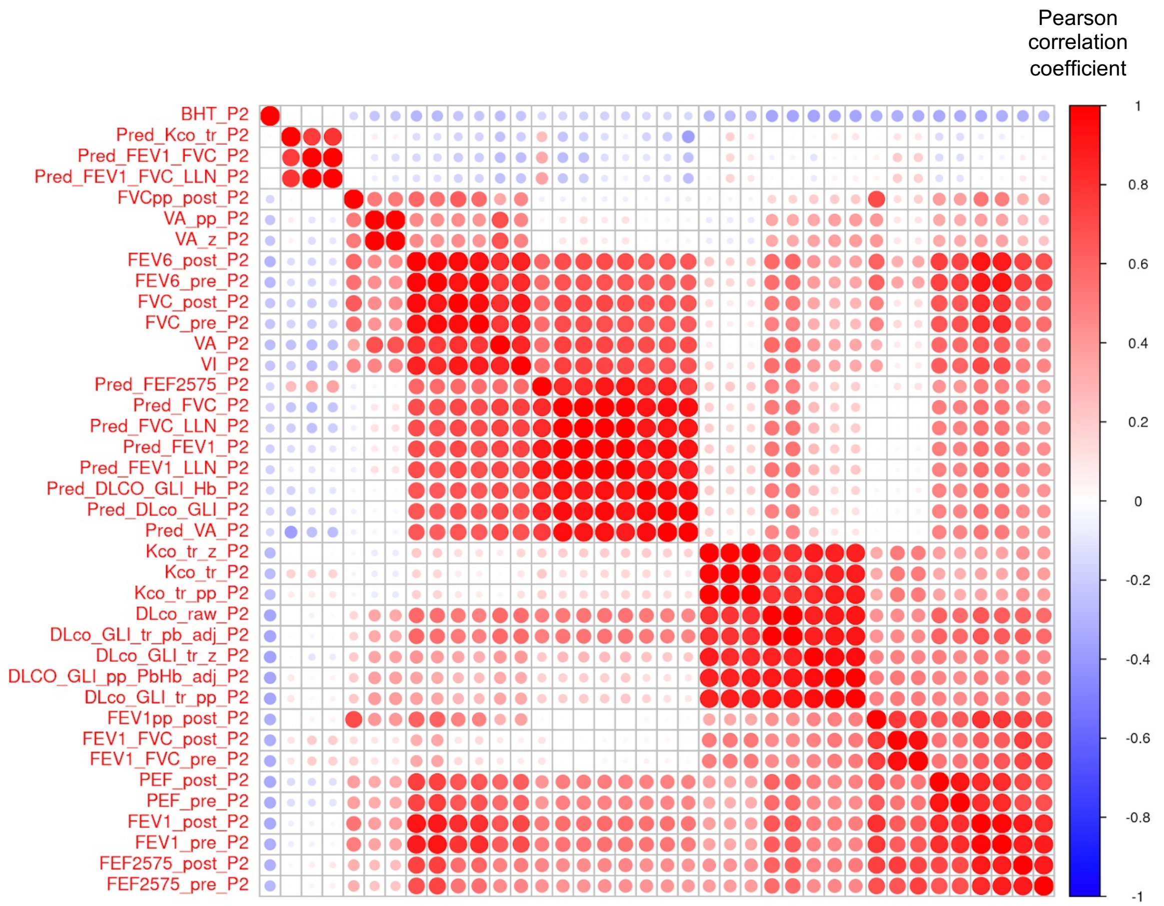
Pearson correlation coefficient
BHT_P2
Pred_Kco_tr_P2
Pred_FEV1_FVC_P2
Pred_FEV1_FVC_LLN_P2
FVCpp_post_P2
VA_pp_P2
VA_z_P2
FEV6_post_P2
FEV6_pre_P2
FVC_post_P2
FVC_pre_P2
VA_P2
VI_P2
Pred_FEF2575_P2
Pred_FVC_P2
Pred_FVC_LLN_P2
Pred_FEV1_P2
Pred_FEV1_LLN_P2
Pred_DLCO_GLI_Hb_P2
Pred_DLco_GLI_P2
Pred_VA_P2
Kco_tr_z_P2
Kco_tr_P2
Kco_tr_pp_P2
DLco_raw_P2
DLco_GLI_tr_pb_adj_P2
DLco_GLI_tr_z_P2
DLCO_GLI_pp_PbHb_adj_P2
DLco_GLI_tr_pp_P2
FEV1pp_post_P2
FEV1_FVC_post_P2
FEV1_FVC_pre_P2
PEF_post_P2
PEF_pre_P2
FEV1_post_P2
FEV1_pre_P2
FEF2575_post_P2
FEF2575_pre_P2
1
0.8
0.6
0.4
0.2
0
-0.2
-0.4
-0.6
-0.8
-1

b

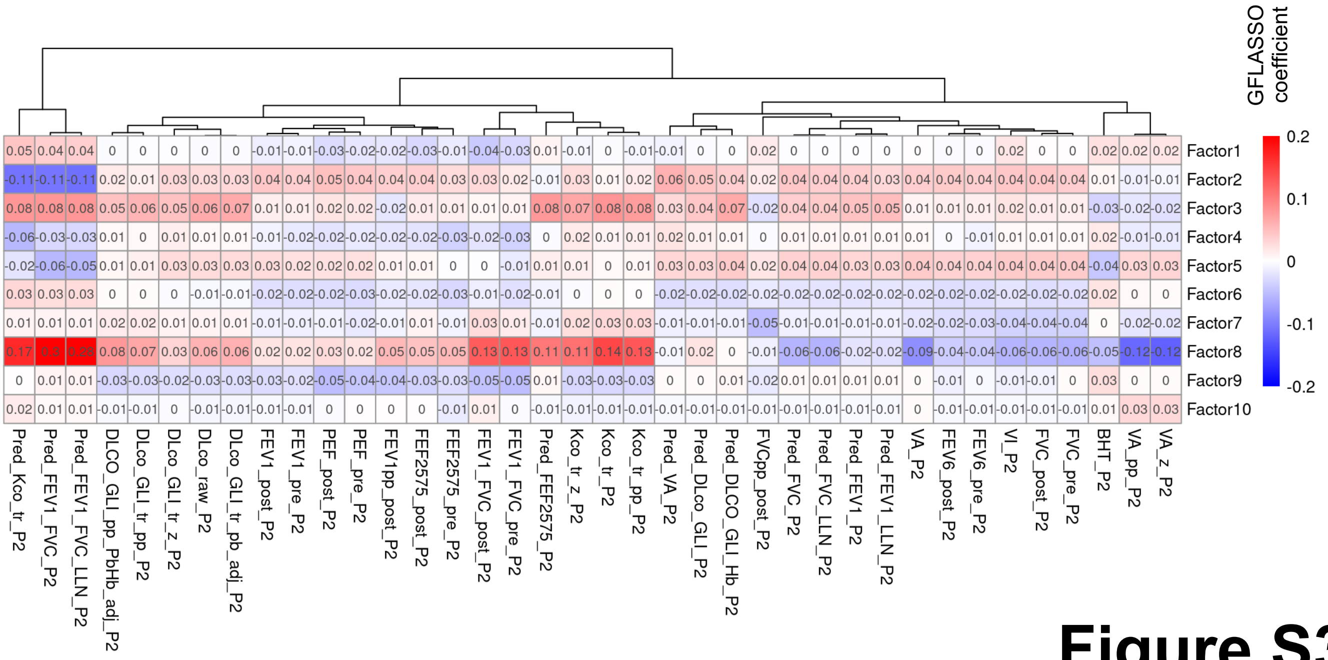
GFLASSO coefficient
0.2
0.1
0
-0.1
-0.2
Factor1
Factor2
Factor3
Factor4
Factor5
Factor6
Factor7
Factor8
Factor9
Factor10
Pred_Kco_tr_P2
Pred_FEV1_FVC_P2
Pred_FEV1_FVC_LLN_P2
DLCO_GLI_pp_PbHb_adj_P2
DLco_GLI_tr_pp_P2
DLco_GLI_tr_z_P2
DLco_raw_P2
DLco_GLI_tr_pb_adj_P2
FEV1_post_P2
FEV1_pre_P2
PEF_post_P2
PEF_pre_P2
FEV1pp_post_P2
FEF2575_post_P2
FEF2575_pre_P2
FEV1_FVC_post_P2
FEV1_FVC_pre_P2
Pred_FEF2575_P2
Kco_tr_z_P2
Kco_tr_P2
Kco_tr_pp_P2
Pred_VA_P2
Pred_DLco_GLI_P2
Pred_DLCO_GLI_Hb_P2
FVCpp_post_P2
Pred_FVC_P2
Pred_FVC_LLN_P2
Pred_FEV1_P2
Pred_FEV1_LLN_P2
VA_P2
FEV6_post_P2
FEV6_pre_P2
VI_P2
FVC_post_P2
FVC_pre_P2
BHT_P2
VA_pp_P2
VA_z_P2

**Figure S3**

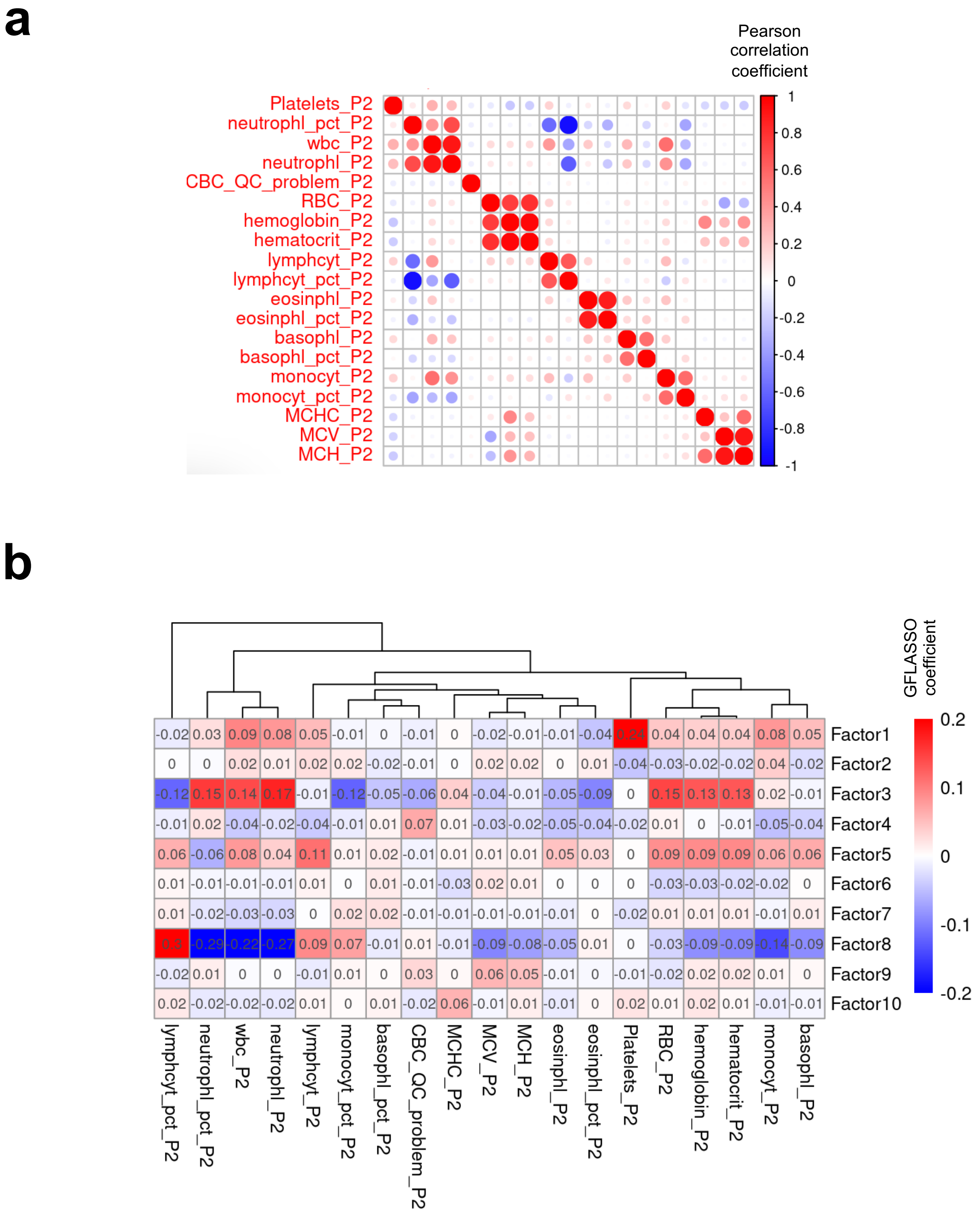
a
Pearson correlation coefficient
1
0.8
0.6
0.4
0.2
0
-0.2
-0.4
-0.6
-0.8
-1
Platelets_P2
neutrophl_pct_P2
wbc_P2
neutrophl_P2
CBC_QC_problem_P2
RBC_P2
hemoglobin_P2
hematocrit_P2
lymphcyt_P2
lymphcyt_pct_P2
eosinphl_P2
eosinphl_pct_P2
basophl_P2
basophl_pct_P2
monocyt_P2
monocyt_pct_P2
MCHC_P2
MCV_P2
MCH_P2
b
GFLASSO coefficient
0.2
0.1
0
-0.1
-0.2
Factor1 -0.02 0.03 0.09 0.08 0.05 -0.01 0 -0.01 0 -0.02 -0.01 -0.01 -0.04 0.24 0.04 0.04 0.04 0.08 0.05
Factor2 0 0 0.02 0.01 0.02 0.02 -0.02 -0.01 0 0.02 0.02 0 0.01 -0.04 -0.03 -0.02 -0.02 0.04 -0.02
Factor3 -0.12 0.15 0.14 0.17 -0.01 -0.12 -0.05 -0.06 0.04 -0.04 -0.01 -0.05 -0.09 0 0.15 0.13 0.13 0.02 -0.01
Factor4 -0.01 0.02 -0.04 -0.02 -0.04 -0.01 0.01 0.07 0.01 -0.03 -0.02 -0.05 -0.04 -0.02 0.01 0 -0.01 -0.05 -0.04
Factor5 0.06 -0.06 0.08 0.04 0.11 0.01 0.02 -0.01 0.01 0.01 0.01 0.05 0.03 0 0.09 0.09 0.09 0.06 0.06
Factor6 0.01 -0.01 -0.01 -0.01 0.01 0 0.01 -0.01 -0.03 0.02 0.01 0 0 0 -0.03 -0.03 -0.02 -0.02 0
Factor7 0.01 -0.02 -0.03 -0.03 0 0.02 0.02 -0.01 -0.01 -0.01 -0.01 -0.01 0 -0.02 0.01 0.01 0.01 -0.01 0.01
Factor8 0.3 -0.29 -0.22 -0.27 0.09 0.07 -0.01 0.01 -0.01 -0.09 -0.08 -0.05 0.01 0 -0.03 -0.09 -0.09 -0.14 -0.09
Factor9 -0.02 0.01 0 0 -0.01 0.01 0 0.03 0 0.06 0.05 -0.01 0 -0.01 -0.02 0.02 0.02 0.01 0
Factor10 0.02 -0.02 -0.02 -0.02 0.01 0 0.01 -0.02 0.06 -0.01 0.01 -0.01 0 0.02 0.01 0.02 0.01 -0.01 -0.01
lymphcyt_pct_P2
neutrophl_pct_P2
wbc_P2
neutrophl_P2
lymphcyt_P2
monocyt_pct_P2
basophl_pct_P2
CBC_QC_problem_P2
MCHC_P2
MCV_P2
MCH_P2
eosinphl_P2
eosinphl_pct_P2
Platelets_P2
RBC_P2
hemoglobin_P2
hematocrit_P2
monocyt_P2
basophl_P2

**Figure S4**

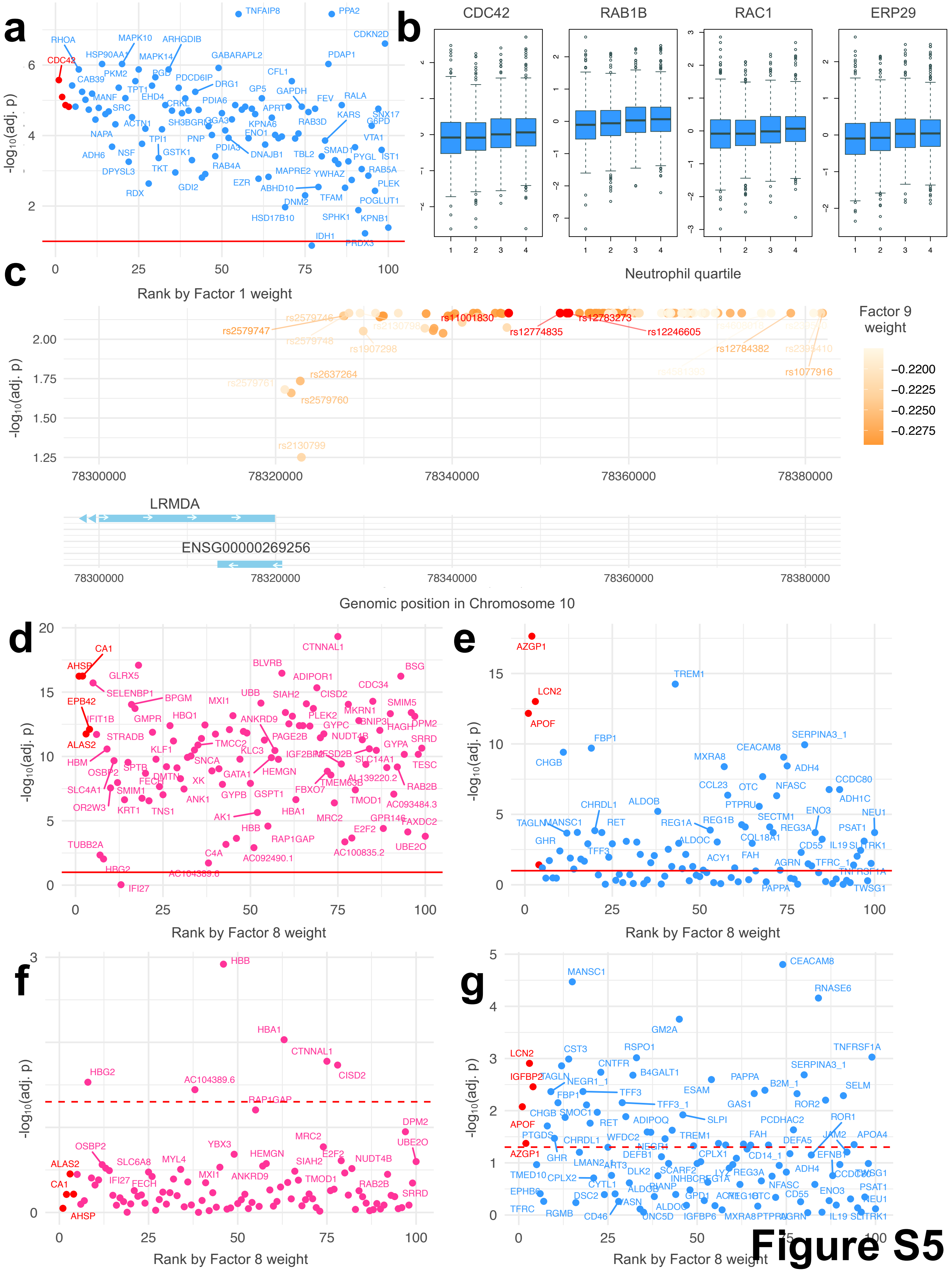
a
b
c
d
e
f
g
-log10(adj. p)
Rank by Factor 1 weight
CDC42
RAB1B
RAC1
ERP29
Neutrophil quartile
Factor 9 weight
Genomic position in Chromosome 10
LRMDA
ENSG00000269256
Rank by Factor 8 weight

Figure S5

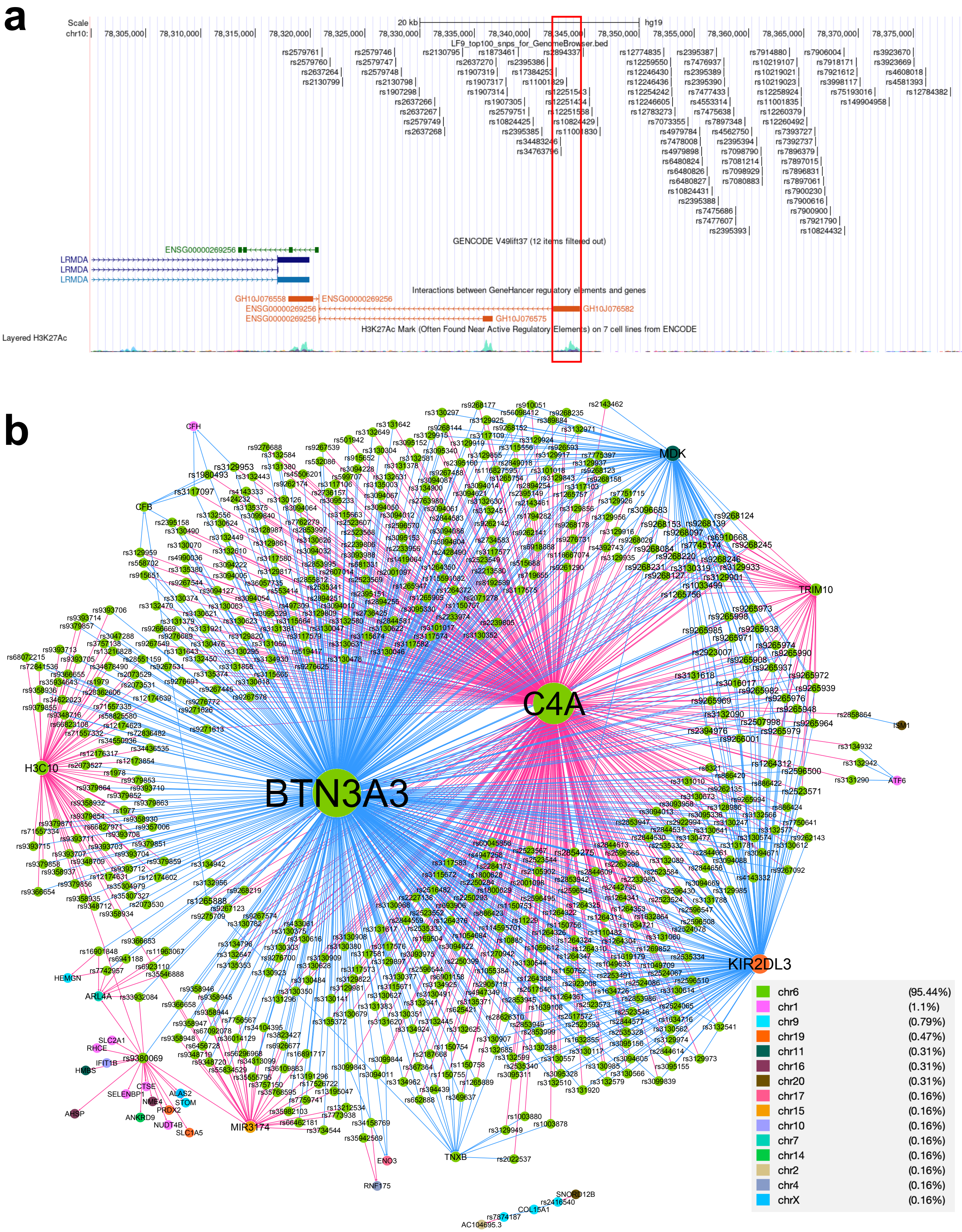
a
Scale
chr10:
20 kb
hg19
LF9_top100_snps_for_GenomeBrowser.bed
GENCODE V49lift37 (12 items filtered out)
ENSG00000269256
LRMDA
LRMDA
LRMDA
Interactions between GeneHancer regulatory elements and genes
GH10J076558
GH10J076582
GH10J076575
H3K27Ac Mark (Often Found Near Active Regulatory Elements) on 7 cell lines from ENCODE
Layered H3K27Ac
b
MDK
TRIM10
C4A
BTN3A3
H3C10
KIR2DL3
TNXB
MIR3174
CFH
CFB
HEMGN
ARL4A
ENO3
RNF175
SNORD12B
COL15A1
AC104695.3
ISM1
ATF6
chr6 (95.44%)
chr1 (1.1%)
chr9 (0.79%)
chr19 (0.47%)
chr11 (0.31%)
chr16 (0.31%)
chr20 (0.31%)
chr17 (0.16%)
chr15 (0.16%)
chr10 (0.16%)
chr7 (0.16%)
chr14 (0.16%)
chr2 (0.16%)
chr4 (0.16%)
chrX (0.16%)

**Figure S6**
